# Genome-wide association study of varenicline-aided smoking cessation

**DOI:** 10.1101/2024.11.13.24317234

**Authors:** Kayesha Coley, Qingning Wang, Richard Packer, Catherine John, Erik Abner, Kadri Reis, Estonian Biobank Research Team, Khaled F. Bedair, Sundararajan Srinivasan, Sara Paciga, Craig Hyde, Robert C. Free, Nicola F. Reeve, David J. Shepherd, Tõnu Esko, Colin Palmer, Ewan Pearson, Anders Malarstig, Martin D. Tobin, Chiara Batini

## Abstract

**Introduction:** Varenicline is an α_4_β_2_ nicotinic acetylcholine receptor partial agonist with the highest therapeutic efficacy of any pharmacological smoking cessation aid and a 12-month cessation rate of 26%. Genetic variation may be associated with varenicline response, but to date no genome-wide association studies of varenicline response have been published.

**Methods:** In this study, we investigated the genetic contribution to varenicline effectiveness using two electronic health record-derived phenotypes. We defined short-term varenicline effectiveness (SVE) and long-term varenicline effectiveness (LVE) by assessing smoking status at 3 and 12 months, respectively, after initiating varenicline treatment. In Stage 1, comprising five European cohort studies, we tested genome-wide associations with SVE (1,405 cases, 2,074 controls) and LVE (1,576 cases, 2,555 controls), defining sentinel variants (the most strongly associated variant within 1 megabase) with *p*-value <5×10^−6^ to follow up in Stage 2. In Stage 2, we tested association between sentinel variants and comparable smoking cessation endpoints in varenicline randomised controlled trials. We subsequently meta-analysed Stages 1 and 2.

**Results:** No variants reached genome-wide significance in the meta-analysis. In Stage 1, 10 sentinel variants were associated with SVE and five with LVE at a suggestive significance threshold (*p*-value <5×10^−6^). None of these sentinels were previously implicated in varenicline-aided smoking cessation or in genetic studies of smoking behaviour.

**Conclusions:** We provide initial insights into the biological underpinnings of varenicline-aided smoking cessation, through implicating genes involved in various processes, including gene expression, cilium assembly and early-stage development.

**Implications:** Leveraging electronic health records, we undertook the largest genetic study of varenicline-aided smoking cessation to date, and the only such study to test genome-wide associations. We showed distinct genetic variants associated (*p*-value <5×10^−6^) with varenicline-aided smoking cessation which implicate diverse cellular functions, including transcriptional regulation, RNA modification and cilium assembly. These provide insights which, if independently corroborated, will improve understanding of varenicline response. The growing availability of biobank resources with genetic and varenicline response data will provide future opportunities for larger studies using the approach we developed.

## Introduction

Tobacco smoking is a major risk factor for many diseases including ischemic heart disease, stroke, chronic obstructive pulmonary disease, and lung cancer^1,2^. Consequently, smoking is a leading cause of preventable morbidity and mortality in the UK^3^, and results in the premature death of over 74,000 smokers each year in England alone^4^. Despite the consistent decline in smoking prevalence in the UK in the last decade, the latest statistics on smoking report that 12.9% of the adult population were smokers in 2022, equating to approximately 6.4 million people^5^.

The substantial impact of smoking on health outcomes has highlighted the importance of quit attempts^2,6^, less than 5% of which are successful without any behavioural support or pharmacological intervention^6^. Nicotine replacement therapy (NRT) and varenicline (Champix) have been prescribed as smoking cessation pharmacotherapies in the UK, with NRT also available over-the counter^7^.

Randomised controlled trials (RCTs) have shown the superiority of varenicline compared to NRT monotherapy for smoking cessation up to 12 months (OR [95% CI] = 1.57 [1.29, 1.91])^8^, and the largest observational study based on electronic health records (EHRs) reported 12-month smoking cessation rates of 26.3% for varenicline and 21.2% for NRT (OR [95% CI] = 1.34 [1.31, 1.38])^9^. The number of items of varenicline dispensed in England has decreased year-on-year since 2010-11, with 232,000 items being dispensed in 2020-21^10^.

Pharmacogenomic studies aim to identify genetic variants that influence how individuals respond to medications, helping to elucidate the mechanisms of drug activity and the reasons for varying effects and adverse reactions among individuals^11^. Varenicline is an α_4_β_2_ nicotinic acetylcholine receptor (nAChR) partial agonist which stimulates the release of dopamine in the brain, thus reducing nicotine cravings and withdrawal symptoms, while promoting smoking abstinence^12^. Several candidate gene association studies, albeit with modest sample sizes, have been performed to investigate the contribution of genetic variation to the success of varenicline-aided quit attempts, and have reported associations with variants in genes encoding nAChR subunits, including *CHRNA4, CHRNA5, CHRNA7* and *CHRNB2*, as well as drug metabolisers (*CYP2B6*)^13–16^. However, candidate gene studies have been generally underpowered, subject to publication biases and are restricted to known biological pathways. In contrast, genome-wide association studies (GWASs), which survey associations with common genetic variants across the genome, have elucidated new biological knowledge^17^ and are increasingly used to study drug response^18^. Here we present the largest GWASs conducted to identify genetic variants associated with the success of varenicline-aided quit attempts, to provide insights into the biological processes underlying the most efficacious pharmacological smoking cessation therapy. We utilise varenicline prescription events and smoking status records in EHRs to define short-term and long-term varenicline effectiveness across five European cohort studies, and combine these results with analyses undertaken in participants from varenicline RCTs conducted by Pfizer in a 2-stage design.

## Methods

### Stage 1 cohorts

Varenicline effectiveness phenotypes included in Stage 1 analyses were defined in Estonian Biobank^19^, the Extended Cohort for E-health, Environment and DNA (EXCEED) Study^20^, Genetics of Diabetes Audit and Research, Tayside and Scotland (GoDARTS)^21^, Genetics of the NHS Scotland Health Research Register (GoSHARE)^22^ and UK Biobank^23^ (further details in **Supplementary Material**).

### Data extraction and phenotype definitions

Varenicline (Champix or Chantix) prescription events, of any pack type, size or dosage, were extracted from prescription EHRs linked to each Stage 1 cohort. The date of the earliest varenicline prescription recorded was defined as the *index date*. With the exception of Estonian Biobank, smoking status was extracted from clinical EHRs using unambiguous Read codes (**Supplementary Table 1**). In Estonian Biobank, smoking status was captured in questionnaire data collected at baseline, due to the limited availability of smoking status records in linked EHRs. Two binary phenotypes were derived from smoking status following an episode of varenicline treatment; cases responded positively to varenicline and quit smoking (i.e. non-smokers), and controls were continuing smokers. For short-term varenicline effectiveness (SVE), smoking status was inferred using the first record between 12 and 104 weeks after *index date,* while for long-term varenicline effectiveness (LVE) we used the first record after 52 weeks from *index date* (**Supplementary Figure 1**). These phenotypes were designed to be consistent with endpoints from previous RCTs and observational studies i.e. at the end of the 12-week standard treatment course, and at 1 year after starting treatment, while allowing sufficient time to capture smoking status due to the irregularity of general practitioner (GP) visitation and recording.

### Descriptive characteristics

In the Stage 1 cohort studies, for SVE and LVE, statistical tests were conducted to examine the relationship between case-control status and several variables, including age (at *index date*), sex, mean number of varenicline prescriptions during a 12-week treatment episode after *index date* (UK Biobank and EXCEED only) and Heaviness of Smoking Index (HSI)^24^ [UK Biobank only]. For categorical variables (sex and HSI), a Chi-square test was used. For continuous variables (age and number of varenicline prescriptions), t-tests were performed.

In UK Biobank, the HSI^24^ was calculated for SVE and LVE cases and controls. HSI ranges from 0 to 6 and is the sum of two categorical variables:

1. Number of cigarettes smoked daily (CPD), where 1-10 is coded as 0, 11-20 as 1, 21-30 as 2, and 31+ as 3.
2. Time from waking to first cigarette (TTFC), where 61+ minutes is coded as 0, 31-60 minutes as 1, 6-30 minutes as 2, and ≤5 minutes as 3.

For both SVE and LVE, we used a Chi-squared test to compare the number of individuals with low-moderate dependence (HSI score ≤4) to those with high dependence (HSI score ≥5) within cases and controls to assess differences in nicotine dependence between these groups.

We also calculated the smoking cessation rate, defined as the proportion of individuals whom, after initiating their first varenicline treatment, were defined as a non-smoker, based on the first smoking status record within the outcome time-period for each phenotype definition. The Clopper-Pearson exact test based on the binomial distribution was used to calculate the 95% confidence interval.

### Power calculations

We calculated power for conducting GWASs of SVE and LVE utilising cases and controls from the Stage 1 cohorts using the Genetic Association Study (GAS) Power Calculator^25^. These calculations were based on an additive genetic model, and trait prevalence was defined as the proportion of cases among the total sample size (**Supplementary Figure 2**).

### Genome-wide association studies

The 2-stage study design is outlined in **Figure 1**. Genotyping quality control and imputation was performed by each individual cohort following the parameters described in **Supplementary Table 2A.** Within each Stage 1 cohort, a GWAS of each varenicline effectiveness phenotype was performed (cohort-specific methodologies are described in **Supplementary Table 2A**). All analyses included only individuals of European ancestry and utilised imputed genomic data for variants with imputation quality ≥0.3, minor allele frequency (MAF) ≥1% and minor allele count (MAC) ≥20. All association testing models utilised imputed dosages and assumed an additive genetic model. Covariates included age (at *index date*), age^2^, sex, up to 10 principal components of genetic ancestry and genotyping array (UK Biobank only). Individuals in GoDARTS genotyped on different arrays were analysed separately and subsequently meta-analysed. All summary statistics were harmonised to genome build hg19 using *liftover*^26^. For each phenotype, GWAS summary statistics from each Stage 1 cohort were meta-analysed using a fixed effect inverse variance weighted model implemented in METAL^27^. Genomic control correction was applied. Only variants represented by either UK Biobank only, or at least two studies were included. Using the same methodology, we also performed a sensitivity analysis excluding GoDARTS from Stage 1.

**Figure 1.**
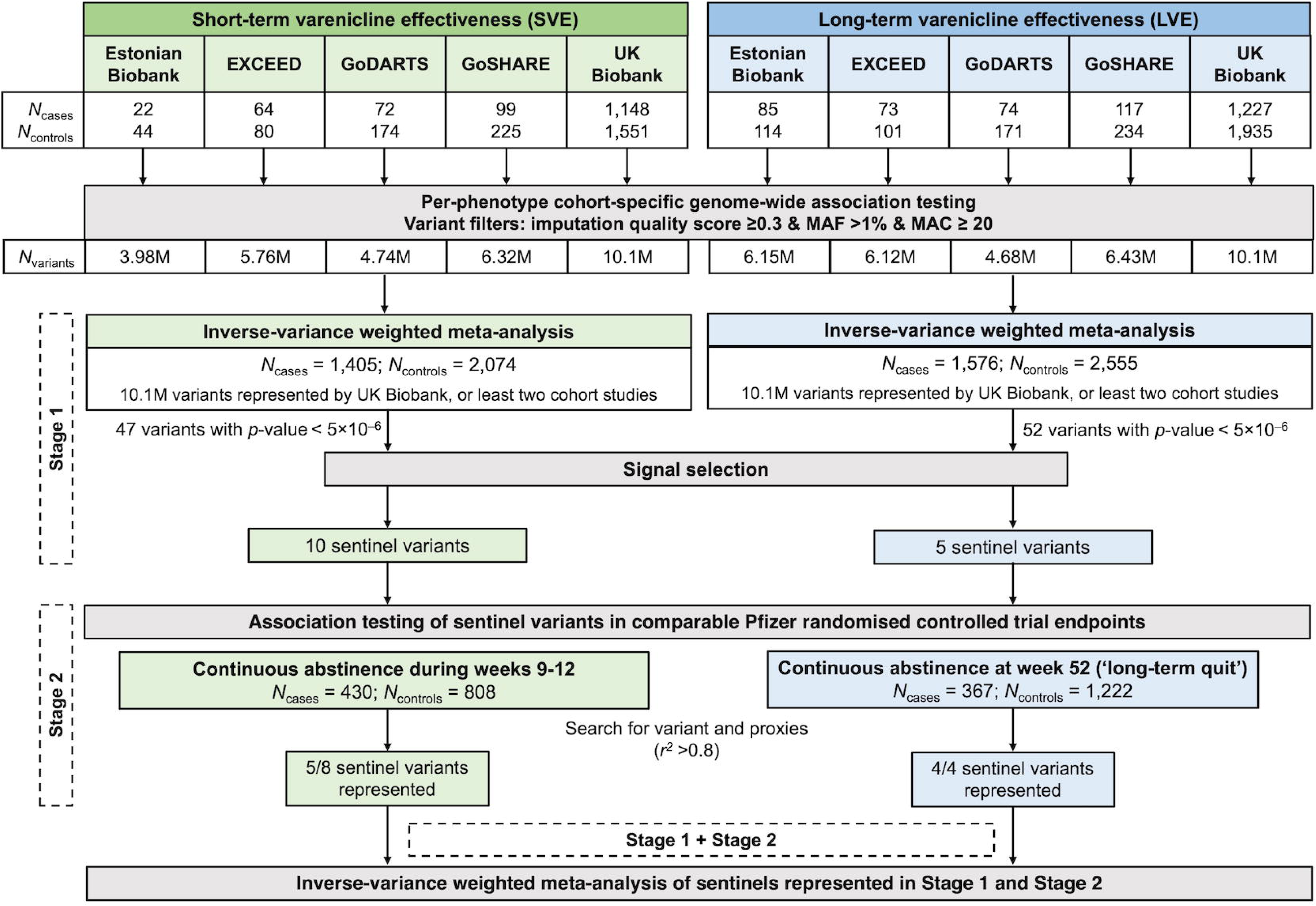
Overview of study design. *Abbreviations:* MAC, minor allele count; MAF, minor allele frequency.

### Sentinel selection

We defined sentinel variants using an iterative procedure by selecting the variant with the lowest *p*- value in Stage 1, excluding all variants within a ±1 megabase (Mb) region around it, and repeating this step until no variants with a *p*-value below the suggestive significance threshold (*p*-value <5×10^−6^) remained. Open Targets Genetics^28^ was used to determine the variant consequence and the gene with the highest ‘overall Variant-to-Gene (V2G)’ score. For variants which were not present in the Open Targets Genetics database, Variant Effect Predictor^29^ was used to define the consequence and ANNOVAR^30^ was used to determine the nearest gene. The biological functions of these genes were investigated using online databases (specifically, Open Targets Platform^31^ and GeneCards^32^) and literature searches.

### Fine-mapping

We fine-mapped each associated locus using the approximate Bayes Factor^33^ method to calculate the posterior probability of inclusion (PIP) for each variant. We set the prior *W* to 0.04 and generated 95% credible sets for each locus by ordering variants by descending PIP, and selecting variants until the until the cumulative PIP was ≥95%.

### Stage 2 dataset

Stage 2 included studies conducted by Pfizer utilising genetic and phenotypic data collected from nicotine-dependent individuals of European genetic ancestry who participated in RCTs assessing varenicline-related endpoints. Genotyping quality control and imputation was performed by Pfizer following the parameters described in **Supplementary Table 2B**. Following the methods described in **Supplementary Table 2B**, sentinel variants from Stage 1 were tested for association with the most comparable RCT smoking cessation endpoint: (i) for SVE, continuous abstinence during weeks 9-12, where cases were defined non-smokers between weeks 9-12 (*N_cases_* = 430) and controls were defined as smokers with a positive carbon monoxide (CO) test at any point between weeks 9-12 (*N_controls_*= 808); and (ii) for LVE, continuous abstinence at week 52 (‘long-term quit’), where cases were defined non-smokers between weeks 9-52 (*N_cases_*= 367), and controls were defined individuals who smoked at any point between weeks 9-52 (*N_controls_* = 1,222). Where sentinels were not present in the Stage 2 genomic data, proxies within ±100 kilobase (kb) in high linkage disequilibrium (LD) [*r*^2^ ≥0.9] were sought using PLINK^34^. Stage 1 and Stage 2 results were meta-analysed using a fixed effect inverse variance weighted model implemented in METAL^27^.

### Comparison to published genetic studies

We performed two look-ups in the Stage 1 GWASs of SVE and LVE, including: (i) variants associated with varenicline efficacy and side effects reported in candidate gene association studies^13–16^; and (ii) independent variants associated with smoking cessation (*p*-value <5×10^−8^) identified in the largest genomic study of smoking cessation to date in European individuals, conducted by the GWAS & Sequencing Consortium of Alcohol and Nicotine use (GSCAN)^35^. Additionally, a look-up of the Stage 1 sentinels (*p*-value <5×10^−6^) associated with SVE and LVE was performed in the European-only results of all the smoking behaviour phenotypes in the most recent GSCAN study^35^; the results used for these look-up analyses were GWASs of smoking initiation, smoking cessation, CPD, and age at smoking initiation. Open Targets Genetics^28^ was also used to determine whether any of the sentinel variants were also associated with other traits analysed in UK Biobank, FinnGen and the GWAS Catalog using the phenome-wide association approach in its ‘PheWAS’ module. The significance level for the PheWAS in Open Targets Genetics is defined after Bonferroni correction for the number of phenotypes tested, considering each as independent^28^.

## Results

### Cohort characteristics

By leveraging varenicline prescription events and post-treatment smoking status records from clinical EHRs or baseline questionnaire data (Estonian Biobank only), we defined two phenotypes: short-term varenicline effectiveness (SVE) and long-term varenicline effectiveness (LVE) [**Supplementary Figure 1**]. **Table 1** outlines the characteristics of cases (successful quitters) and controls (continuing smokers) identified in each Stage 1 cohort (Estonian Biobank^19^, EXCEED^20^, GoDARTS^21^, GoSHARE^22^ and UK Biobank^23^) for inclusion in GWASs of each phenotype. Across both phenotypes defined in the five studies, there was no difference in the age of cases and controls, and only two studies (Estonian Biobank and GoDARTS) showed significant differences in the proportion of female cases compared to controls for SVE. For SVE, the smoking cessation rate ranged from 29.3% in GoDARTS to 44.4% in EXCEED, and for LVE, the smoking cessation rate was highest in Estonian Biobank at 42.7% and lowest in GoDARTS at 30.2%. GoDARTS had the lowest cessation rates in the short-term and long-term, suggesting barriers to sustained smoking abstinence, or an effect of the cohort recruitment strategy, whereby approximately half of the cohort have type 2 diabetes, for which smoking is an important risk factor^36^. Interrogation of individual-level data in UK Biobank and EXCEED showed that, for both phenotypes, controls had fewer varenicline prescriptions within the 12-week treatment period than cases. Additionally, for both phenotypes defined in UK Biobank, there were more individuals with high nicotine dependence (corresponding to a HSI^24^ score of 5 or 6) in the control group, compared to cases (*p*-value <0.001). For SVE, 13.3% of controls compared to 6.0% of cases had high nicotine dependence, while for LVE, 11.8% of controls compared to 6.8% of cases had high nicotine dependence.

**Table 1.**
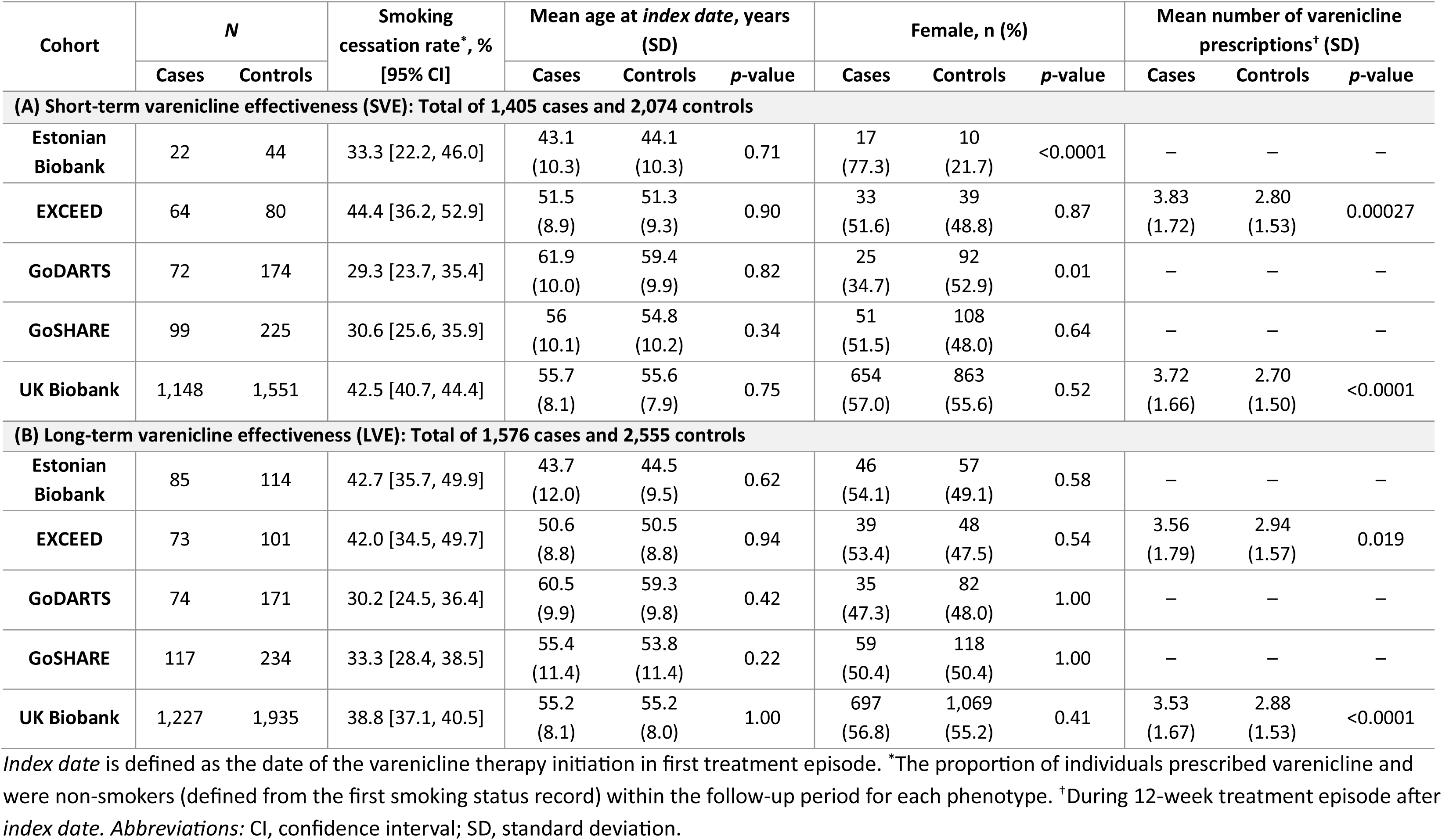
Case-control characteristics for (A) SVE and (B) LVE defined in Stage 1 cohorts.

### Genome-wide association studies of varenicline-aided smoking cessation

For both varenicline effectiveness phenotypes, we conducted GWASs in each Stage 1 cohort and meta-analysed the results. **Figure 1** outlines the number of individuals and variants included in each analysis stage. The Stage 1 GWAS included up to 3,479 individuals (1,405 cases and 2,074 controls) for SVE, and up to 4,131 individuals (1,576 cases and 2,555 controls) for LVE. Both Stage 1 GWASs included over 10 million variants and did not show any evidence of test statistic inflation with λ_GC_ estimates of 0.996 and 0.994 for SVE and LVE, respectively (**Figure 2**; **Supplementary Figure 3**). We identified 10 sentinel variants associated with SVE and five sentinel variants associated with LVE that reached our pre-defined statistical threshold (*p*-value <5×10^−6^) [**Table 2**; further details in **Supplementary Table 3**]. There was no overlap (considering an LD threshold of *r*^2^ >0.8) between the sentinel variants for SVE and the sentinel variants for LVE. We compared effect size estimates for the sentinel variants after the exclusion of GoDARTS (as 50% of GoDARTS participants had type 2 diabetes) and found that this had no impact (**Supplementary Figure 4**). The 95% credible set for each locus (within which we expect the causal variant to lie with 95% probability) contained a median of 9 variants (range 1-105 variants) [**Supplementary Table 4**]. One locus (represented by sentinel rs2297038) had a single putative causal variant, and eight loci (represented by sentinel variants rs4364036, rs4686373, rs78597169, rs2297038, rs149318872, rs10421326, rs35443123 and rs766169) had a single variant with PIP >50%, all of which were also the sentinel variant.

**Figure 2.**
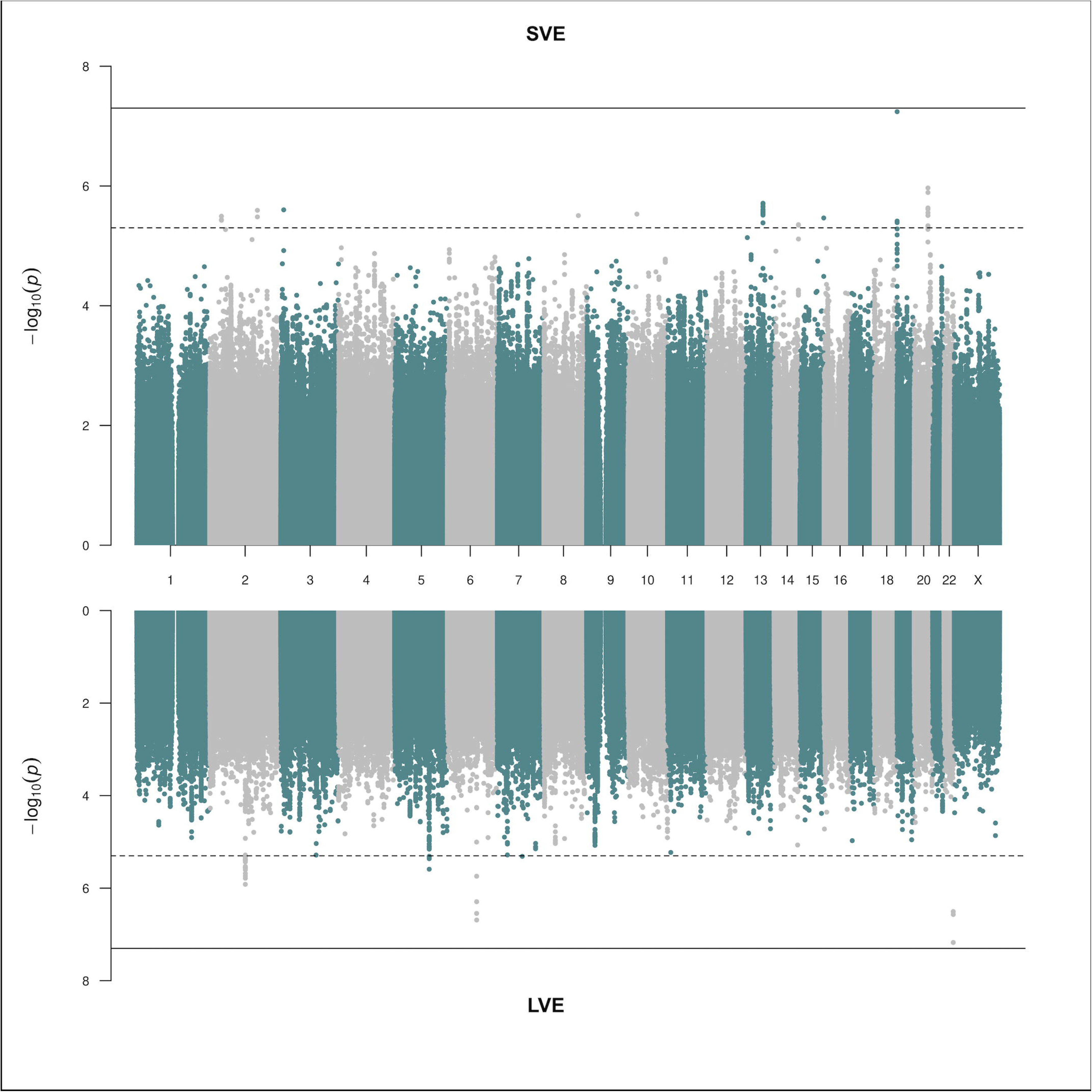
Miami plot for Stage 1 GWASs of short-term varenicline effectiveness (SVE) and long-term varenicline effectiveness (LVE). The grey solid line represents the genome-wide significance threshold (*p*-value <5×10^−8^), and the grey dotted line represents the suggestive significance threshold (*p*-value <5×10^−6^).

**Table 2.**
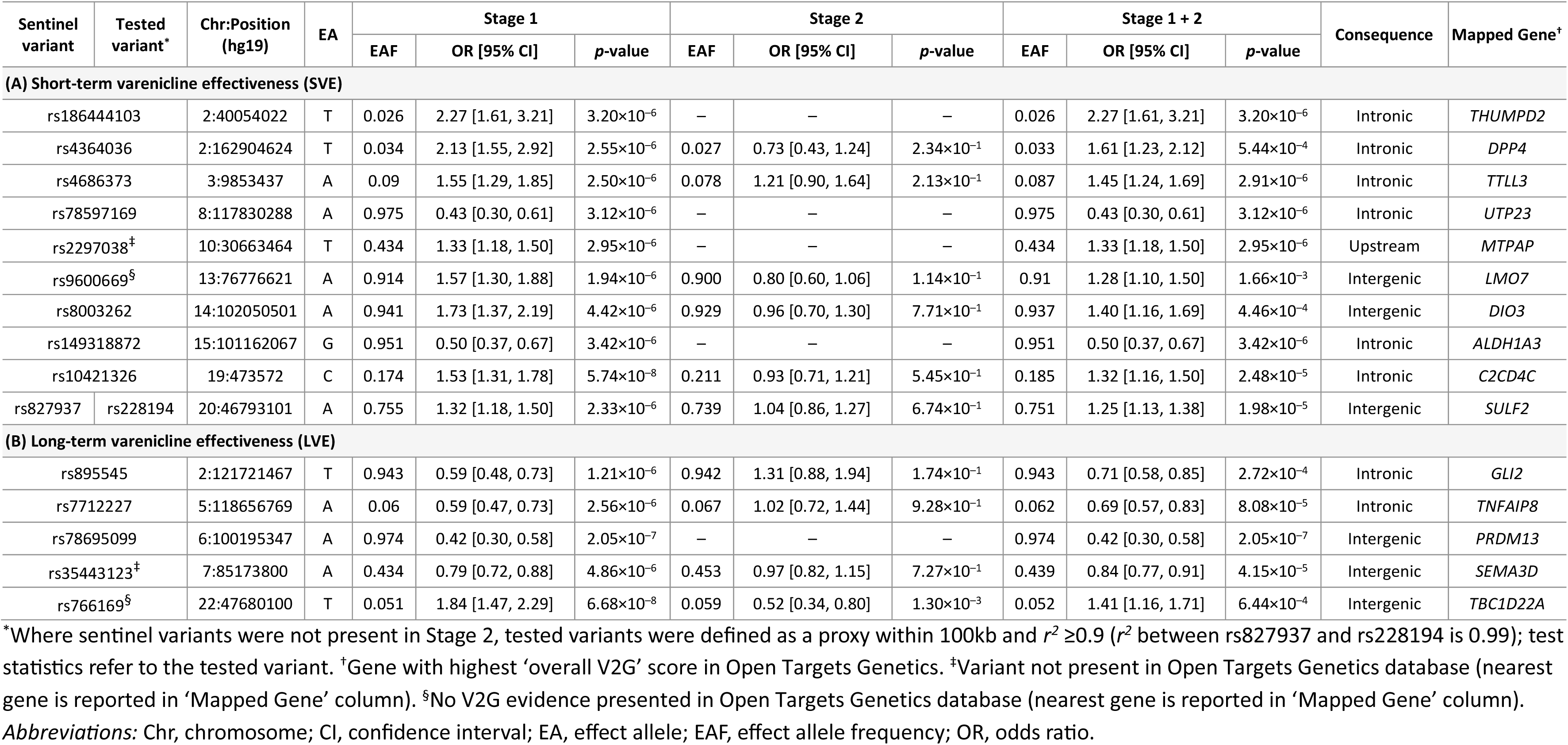
Sentinel variants associated with (A) SVE and (B) LVE in Stage 1 GWASs.

The Stage 2 dataset comprised 430 cases and 808 controls for SVE and 367 cases and 1,222 controls for LVE from RCTs. Ten of the 15 sentinel variants (six SVE sentinel variants and four LVE sentinel variants) from Stage 1 were available for follow up in Stage 2 (**Figure 1**; **Table 2**). As five of the 15 Stage 1 sentinel variants were not represented in Stage 2 and as no variants reached genome-wide significance (*p*-value <5×10^−8^) in the meta-analysis of Stages 1 and 2 (**Table 2**), we describe all 15 variants taken into Stage 2 below (that is, reaching *p*-value <5×10^−6^ in Stage 1). These 15 sentinels collectively implicated 15 genes: *ALDH1A3*, *C2CD4C*, *DIO3*, *DPP4*, *LMO7*, *MTPAP*, *SULF2*, *THUMPD2*, *TTLL3* and *UTP23* for SVE, and *GLI2*, *PRDM13*, *SEMA3D*, *TBC1D22A* and *TNFAIP8* for LVE (**Table 2**).

These genes are involved in the regulation of transcription (*GLI2* and *LMO7),* chromatin remodelling and methylation (*PRDM13*), processing of messenger RNA (*MTPAP*) and ribosomal RNA (*UTP23*), and transfer RNA methylation (*THUMPD2*)^31,32^. Additionally, some genes are essential for normal fetal development, including *ALDH1A3* for eye development, *DIO3* for thyroid hormone level regulation and *SEMA3D* for axon guidance and cell migration^31,32^. Further gene functions include the regulation of apoptosis (*TNFAIP8*), glycoprotein metabolism (*SULF2*), glucose and insulin metabolism (*DPP4*), and cilium assembly (*TTLL3* and *TBC1D22A*)^31,32^. *TBC1D22A* is also likely to be involved in intracellular protein transport and GTPase activation^32^.

### Phenome-wide association studies and comparison to published genetic studies

Using the repository of summary statistics from UK Biobank, FinnGen and the GWAS Catalog present in Open Targets Genetics^28^, phenome-wide association studies (PheWASs) showed that two of the sentinels (*C2CD4C*-rs10421326 and *TNFAIP8*-rs7712227) were associated with various blood cell measurements. rs10421326-C (associated with increased varenicline effectiveness), was associated with increased lymphocyte count, reticulocyte count and related cellular phenotypes, while rs7712227-G (associated with increased varenicline effectiveness) was significantly associated with reduced mean reticulocyte volume.

Of the variants previously associated with varenicline efficacy in candidate gene studies^13–16^, only one reached a Bonferroni corrected threshold of *p*-value <8.33×10^−3^ (based on six independent tests considering variants as independent using an LD threshold of *r*^2^ <0.2) in our Stage 1 GWAS of LVE (**Supplementary Table 5**). However, this variant, *CHRNA4*-rs2236196, which was previously associated with continuous abstinence at weeks 9-12 after varenicline treatment (OR [95% CI] = 1.54 [1.13, 2.09])^13^, had an opposite direction of effect to the LVE GWAS (OR [95% CI] = 0.86 [0.77, 0.96]) for allele G. Further, none of the variants associated with either nausea severity post-varenicline treatment in candidate gene association studies, or smoking cessation in GWAS, were associated with either SVE or LVE at a Bonferroni corrected threshold based on the number of independent tests (LD threshold of *r*^2^ <0.2) across each phenotype (**Supplementary Table 5**).

We determined whether any of our varenicline-mediated smoking cessation sentinel variants were previously associated with smoking behaviours (**Supplementary Table 6**). The LVE sentinel variant *PRDM13*-rs78695099 (not available to study in Stage 2) was associated with smoking behaviours, specifically smoking initiation (*p*-value = 0.00168), at a Bonferroni corrected *p*-value threshold based on 15 independent tests. Allele A of rs78695099, associated with a lower OR of quitting after varenicline treatment, conferred a reduced risk of smoking initiation.

## Discussion

In this study, we leveraged routinely collected health data to undertake GWASs of varenicline-aided smoking cessation. Fifteen sentinel variants met our pre-defined statistical threshold (*p*-value <5×10^−6^) for SVE or LVE in Stage 1 (**Table 2**; **Supplementary Table 3**), and no overlap was observed between the variants associated with the two traits. None of these sentinels have been previously implicated in candidate gene association studies of varenicline-aided smoking cessation or in GWASs of smoking behaviour, and variants previously reported to be associated with varenicline efficacy, nausea, or smoking cessation were not associated with varenicline-aided smoking cessation in our study (**Supplementary Table 5**).

The 15 sentinels implicate genes that, individually, exhibit diverse cellular functions, with many involved in shared biological processes, including gene expression, specifically transcriptional regulation (*GLI2, LMO7* and *PRDM13*) and RNA modification (*MTPAP*, *UTP23*, and *THUMPD2*), cilium assembly (*TTLL3* and *TBC1D22A*), as well as processes essential for fetal growth and development (*ALDH1A3, DIO3*, *GLI2 and SEMA3D*)^31,32^. Further, *TBC1D22A,* which was implicated by LVE sentinel variant rs766169, has also been previously reported in smoking cessation by a statistically independent variant^35^. With the caveat that future replication would be required, our investigations into the functions of the genes implicated by these variants provides initial insights into potential biological mechanisms.

Individuals who continue smoking after varenicline treatment may do so for various reasons beyond those relating to pharmacological efficacy, such as nicotine dependence or non-adherence potentially due to adverse reactions. We observed that controls (i.e. continuing smokers) had significantly higher nicotine dependence and received significantly fewer varenicline prescriptions, compared to cases. This suggests that non-adherence to treatment in the control group could be explained by lack of motivation or adverse reactions, thereby impacting on varenicline continuation and successful quit attempts.

By harnessing EHRs linked to five cohort studies, we have defined varenicline effectiveness with sample sizes almost 8-times larger than the largest candidate gene study of varenicline-aided smoking cessation^13^. To assess effectiveness, we considered only the first treatment episode to ensure a consistent approach to the clinical question being addressed by the study. Another strength of this study is the use of an unbiased GWAS approach, which allowed us to study over 10 million variants across the genome and identify novel associations. Previously, pharmacogenetic studies of this phenotype have taken a candidate gene approach where associations are limited to several hundred variants across a predefined set of likely implicated genes^13–16^.

Our study was well powered to detect variant associations of relatively large effect. For example, the study was well-powered at a genome-wide significance threshold (*p*-value <5×10^−8^) to detect common variants (MAF >40%) with an odds ratio as low as 1.2 for both varenicline response phenotypes examined (**Supplementary Figure 2**). Whilst larger effect size estimates have been reported for some genetic determinants of drug response^37^, common genetic variants associated with common complex traits have been typically more modest^17^. Replicable genetic associations of common complex traits have typically emerged in small numbers once several thousand cases were studied, and replicable associations increased dramatically once tens of thousands of cases became available to study^17^. Genetic studies of drug response have typically been much smaller, and whilst larger than previous candidate gene association studies of varenicline response^13–16^, fewer than 1,600 cases were available in each of our stages. In this context, the lack of replicable findings to date indicates the need for larger sample sizes. For studies utilising EHR-based phenotypes, this could be achieved through a combination of further data linkage in participants who have already consented to this (for example, UK Biobank so far has linked primary care EHRs on less than one-half of consenting participants) and through additional Biobanks (such as *All of Us*^38^ and Our Future Health^39^) as data becomes available. Denser genotyping arrays and new imputations would improve the availability of data for follow up; this is important to address given the unavailability of five of the 15 sentinels in Stage 2. Our Stage 1 studies were largely population-based studies or biobanks with healthy participants. This would be expected to impact on power where there is a lower proportion of smoking and could also impact on generalisability of findings to those with disease (although effect estimates did not alter when GoDARTS was excluded, in which 50% of participants had type 2 diabetes). Further, these analyses were restricted to individuals of European genetic ancestry and utilising similar data from developing initiatives with focused recruitment in minority ethnic populations^38,39^ will help to ensure the generalisability of findings. In summary, despite being the largest global study varenicline response, the associations we discovered did not reach genome-wide significance (*p*-value <5×10^−8^) and need to be viewed with caution pending independent replication or larger meta-analyses that corroborate these findings. Factors impacting on the power to detect genetic associations at this threshold include: (i) underlying effect sizes of common variants associated with varenicline response likely to be more modest than originally assumed; (ii) density of genotyping platforms and of imputation panels, such that not all studies contributed to meta-analysis of a given variant; (iii) possible measurement error in defining the phenotype from EHRs.

Varenicline was still being widely prescribed when we initiated this study, though its production was halted in July 2021 due to a nitrosamine impurity^40,41^. In considering the future availability and use of varenicline, we note the following: (i) varenicline is the most efficacious smoking cessation therapy, was approved for use in 116 countries and has been prescribed to 24 million smokers^40^; (ii) the withdrawal of varenicline was not due to concerns about efficacy or side effects, but related to the theoretical risks of cancer attributable to the nitrosamine impurity^40^; (iii) the health benefits of stopping smoking on cancer risk substantially outweigh cancer risk from the nitrosamine impurity^40^; (iv) the Federal Drug Administration (FDA) stated its confidence in the ability to manufacture varenicline containing ≤37ng/day of the N-nitroso-varenicline impurity (the agency’s acceptable intake limit)^42^; (v) varenicline is now off-patent and FDA-approved generic varenicline became available in September 2021, after which a rise in prescriptions was observed in the US^41^ and; (vi) generic varenicline was introduced in the UK in August 2024^43^.

In conclusion, this is largest genome-wide association study of varenicline-aided smoking cessation to date, which incorporates definitions of short- and long-term cessation, and implicates a diverse range of processes, including gene expression, cilium assembly and fetal development. Larger and more diverse populations will be required for powerful, generalisable genetic association studies of varenicline-aided smoking cessation. The EHR-based phenotyping approach we present here can be applied once additional data becomes available in UK Biobank and other large studies under development, facilitating the corroboration of findings from this study.

## Data availability

Summary statistics from SVE and LVE Stage 1 meta-analyses are being deposited at the NHGRI-EBI GWAS Catalog (https://www.ebi.ac.uk/gwas/) and accession numbers will be added as soon as they are available.

## Acknowledgements

We thank all participants of the cohort studies and randomised controlled trials used in this project. This study used the ALICE and SPECTRE High Performance Computing Facilities at the University of Leicester. The Estonian Genome Center analyses were partially carried out in the High Performance Computing Center, University of Tartu.

**Estonian Biobank:** The activities of the Estonian Biobank are regulated by the Human Genes Research Act, which was adopted in 2000 specifically for the operations of Estonian Biobank.

Individual level data analysis in Estonian Biobank was carried out under ethical approval 1.1-12/624 from the Estonian Committee on Bioethics and Human Research (Estonian Ministry of Social Affairs), using data according to release application 6-7/GI/33501 from the Estonian Biobank. The work of the Estonian Genome Center, University of Tartu was funded by the European Union through Horizon 2020 research and innovation program under grants no. 810645 and 894987, through the European Regional Development Fund projects GENTRANSMED (2014-2020.4.01.15-0012), MOBEC008, MOBERA21 and the Estonian Research Council grant PUT (PRG1291, PRG687 and PRG184).

**EXCEED:** EXCEED is supported by the University of Leicester, the National Institute for Health and Care Research Leicester Respiratory Biomedical Research Centre, the Wellcome Trust (WT 202849), and Cohort Access fees from studies funded by the Medical Research Council (MRC), Biotechnology and Biological Sciences Research Council, National Institute for Health and Care Research, the UK Space Agency, and GlaxoSmithKline. It was previously supported by Medical Research Council grant G0902313. EXCEED is supported by BREATHE - The Health Data Research Hub for Respiratory Health (UKRI_PC_19004) in partnership with SAIL Databank. BREATHE is funded through the UK Research and Innovation Industrial Strategy Challenge Fund and delivered through Health Data Research UK.

EXCEED received ethical approval from the Leicester Central Research Ethics Committee (13/EM/0226). Substantial amendments have been approved by the same Research Ethics Committee for the collection of new data relating to the COVID-19 pandemic, including the COVID-19 questionnaires and antibody testing.

**GoDARTS & GoSHARE:** We are grateful to all the participants of GoDARTS and SHARE study, the general practitioners, the Scottish School of Primary Care for their help in recruiting the participants and to the whole team, which includes interviewers, computer and laboratory technicians, clerical workers, research scientists, volunteers, managers, receptionists, and nurses. The study complies with the Declaration of Helsinki. We acknowledge the support of the Health Informatics Centre, University of Dundee, for managing and supplying the anonymized data and NHS Tayside, the original data owner. GoDARTS and SHARE studies were reviewed and approved by Tayside Medical Ethics Committee 053/04 and East of Scotland Ethics Committee NHS REC 13/ES/0020. The Wellcome Trust United Kingdom Type 2 Diabetes Case Control Collection (supporting GoDARTS) was funded by the Wellcome Trust (072960/Z/03/Z, 084726/Z/08/Z, 084727/Z/08/Z, 085475/Z/08/Z, 085475/B/08/Z) and as part of the EU IMI-SUMMIT programme. GoSHARE is funded by NHS Research Scotland (NRS) (http://www.nhsresearchscotland.org.uk/), a partnership of Scottish Health Boards and the Scottish Government Chief Scientist Office (CSO) that oversees governance and management of clinical research.

**UK Biobank:** UK Biobank is generously supported by its founding funders the Wellcome Trust and UK Medical Research Council, as well as the Department of Health, Scottish Government, the Northwest Regional Development Agency, British Heart Foundation and Cancer Research UK. The UK Biobank genetic and phenotypic data were obtained under UK Biobank Applications 4982 and 59822. UK Biobank has ethical approval from the UK National Health Service (NHS) National Research Ethics Service (11/NW/0382).

For the purpose of open access, the author has applied a CC BY public copyright licence to any Author Accepted Manuscript version arising from this submission.

