## Supplementary Material for "Genome-wide association study of varenicline-aided smoking cessation"

### Supplementary Text

#### Stage 1 cohorts

Estonian Biobank[1] is a population-based cohort with deep baseline phenotyping and clinical events for over 200,000 participants aged ≥18 years from Estonia. In this study, data freeze (2020v4) was used. All biobank participants have signed a broad informed consent form and information on ICD-10 codes is obtained via regular linking with the national Health Insurance Fund and other relevant databases, with majority of the electronic health records having been collected since 2004.

The Extended Cohort for E-health, Environment and DNA (EXCEED) Study[2] is a cohort of approximately 10,000 participants (aged between 40 and 69 years) recruited from Leicester, Leicestershire and Rutland, mainly through local general practices since 2013.

Genetics of Diabetes Audit and Research in Tayside Scotland (GoDARTS)[3] is a cohort study of over 16,000 individuals, approximately 50% of which have type II diabetes, from the Tayside region of Scotland, and includes lifestyle data, biomarker measurements and medical records.

Genetics of the Scottish Health Research Register (GoSHARE)[4] is a sub-study of the SHARE initiative where almost 75,000 individuals living in Scotland have linked EHRs and consented for genotyping analysis.

UK Biobank[5] is a cohort of approximately 500,000 individuals recruited from across the United Kingdom. Individuals aged between 40 and 69 years were recruited from the general population between 2006 and 2010.

### Supplementary Figures

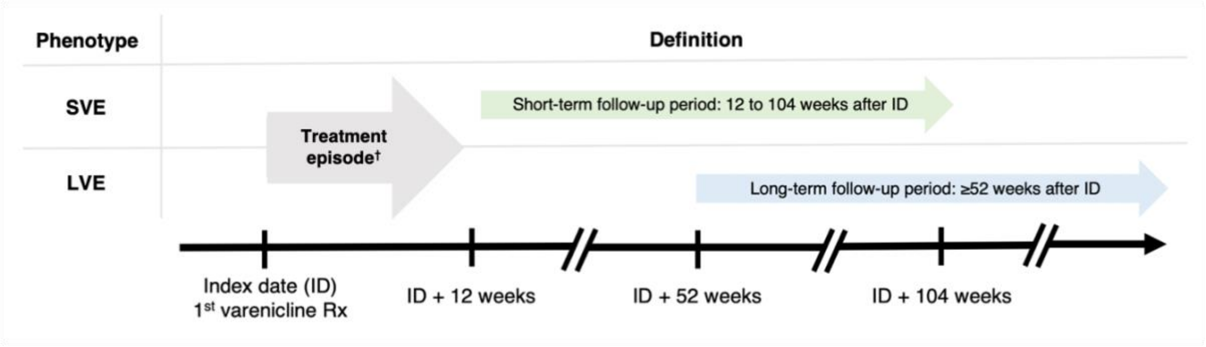

**Supplementary Figure 1** Description of phenotype definitions for SVE and LVE defined in cohort studies. †Based on a standard 12-week treatment plan. *Abbreviations:* Rx, prescription.

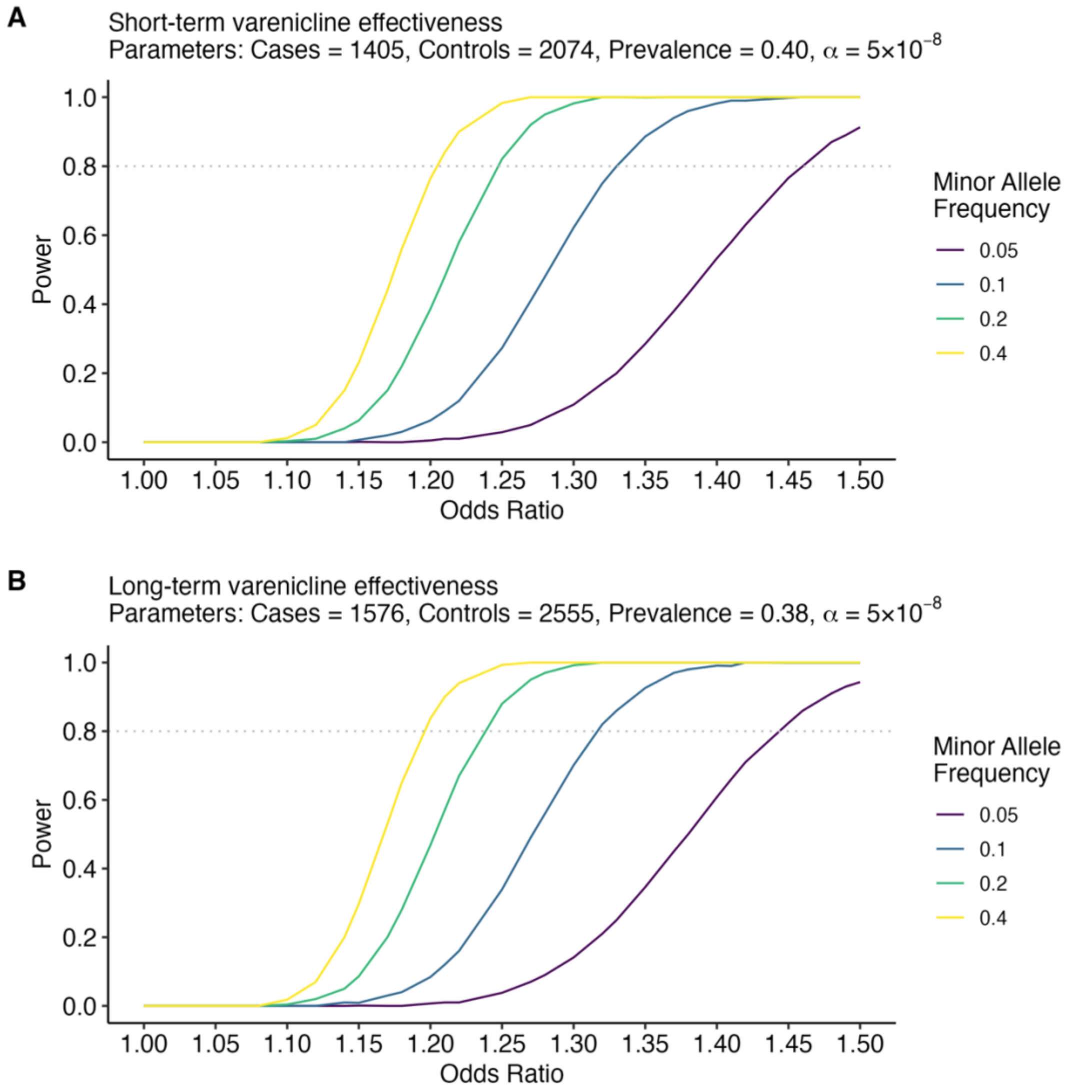

**Supplementary Figure 2** Power-odds ratio curves for Stage 1 GWASs of **(A)** SVE and **(B)** LVE.

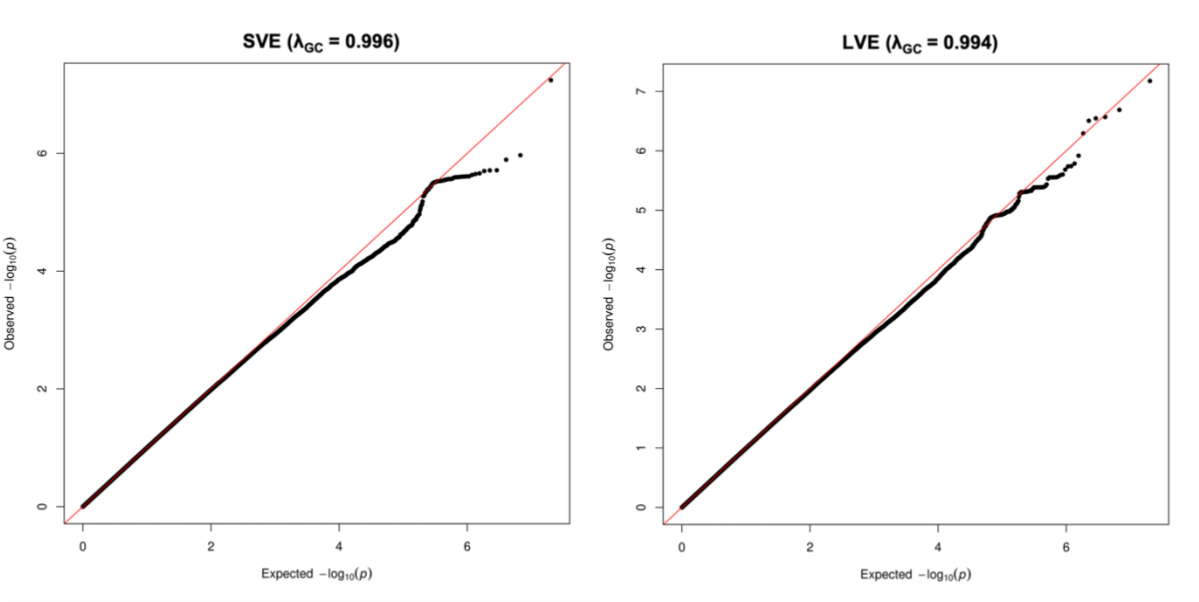

**Supplementary Figure 3** QQ plots for Stage 1 GWASs of short-term varenicline effectiveness (SVE) and long-term varenicline effectiveness (LVE). $\lambda_{GC}$, genomic control lambda.

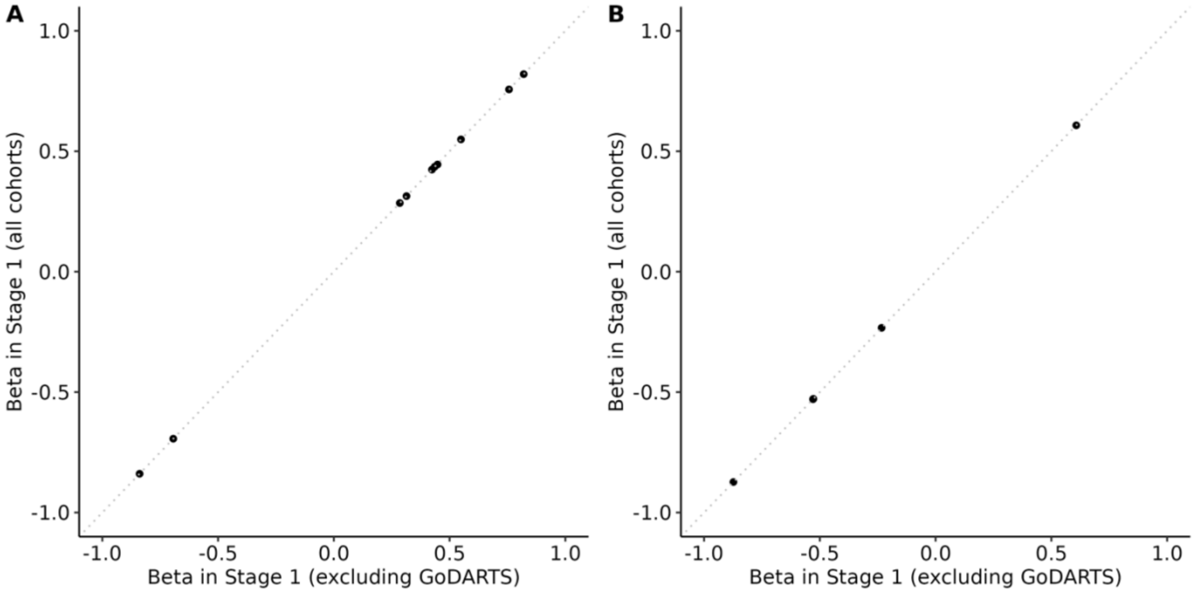

**Supplementary Figure 4** Stage 1 effect size (beta) comparison of **(A)** SVE and **(B)** LVE sentinels when including all cohorts, compared to excluding GoDARTS.

### Supplementary Tables

**Supplementary Table 1** Read v2 and Read v3 (CTV3) codes used to define smoking status.

| Code | | Description | Inferred smoking status |
|---|---|---|---|
| **Read v2** | **Read v3** | | |
| 1371. | XE0oh | Never smoked tobacco | Non-smoker |
| | Ub0oq | Non-smoker | Non-smoker |
| 1372. | XE0oi | Trivial smoker - < 1 cig/day | Smoker |
| | Ub1tR | Occasional cigarette smoker | Smoker |
| 1373. | 1373. | Light smoker - 1-9 cigs/day | Smoker |
| 1374. | 1374. | Moderate smoker - 10-19 cigs/d | Smoker |
| 1375. | 1375. | Heavy smoker - 20-39 cigs/day | Smoker |
| 1376. | 1376. | Very heavy smoker - 40+cigs/d | Smoker |
| 1377. | 1377. | Ex-trivial smoker (<1/day) | Non-smoker |
| 1378. | 1378. | Ex-light smoker (1-9/day) | Non-smoker |
| 1379. | 1379. | Ex-moderate smoker (10-19/day) | Non-smoker |
| 137A. | 137A. | Ex-heavy smoker (20-39/day) | Non-smoker |
| 137B. | 137B. | Ex-very heavy smoker (40+/day) | Non-smoker |
| 137e. | XaBSp | Smoking restarted | Smoker |
| 137F. | 137F. | Ex-smoker - amount unknown | Non-smoker |
| 137H. | 137H. | Pipe smoker | Smoker |
| 137j. | Xa1bv | Ex-cigarette smoker | Non-smoker |
| 137J. | 137J. | Cigar smoker | Smoker |
| 137K. | 137K. | Stopped smoking | Non-Smoker |
| 137K0 | XaQzw | Recently stopped smoking | Non-smoker |
| 137l. | XaQ8V | Ex roll-up cigarette smoker | Non-smoker |
| 137m. | XaWNE | Failed attempt to stop smoking | Smoker |
| 137L. | 137L. | Current non-smoker | Non-smoker |
| 137N. | 137N. | Ex pipe smoker | Non-smoker |
| 137O. | 137O. | Ex cigar smoker | Non-smoker |
| 137P. | XE0oq | Cigarette smoker | Smoker |
| 137R. | 137R. | Current smoker | Smoker |
| 137S. | Ub1na | Ex-smoker | Non-smoker |

**Supplementary Table 2** Characteristics of **(A)** participating cohorts in the Stage 1 GWASs of varenicline effectiveness phenotypes, and **(B)** the Stage 2 dataset of comparable randomised controlled trial (RCT) endpoints.

| | **(A) Stage 1** | | | | | **(B) Stage 2** |
|---|---|---|---|---|---|---|
| **Cohort** | **Estonian Biobank** | **EXCEED** | **GoDARTS** | **GoSHARE** | **UK Biobank** | **Pfizer** |
| **Recruitment country** | Estonia | UK | UK (Scotland) | UK (Scotland) | UK | Worldwide |
| **Phenotype data** | EHRs and questionnaire data* | EHRs | EHRs | EHRs | EHRs | RCTs |
| **Genomic data‡** | | | | | | |
| **Genotyping array** | Illumina Global Screening v1.0 and v2.0 | Applied Biosystems UK Biobank Axiom | Affymetrix SNP 6.0; Illumina HumanOmniExpress; Illumina Infinium Broad† | Illumina Global Screening v2.0 | Applied Biosystems UK BiLEVE Axiom; Applied Biosystems UK Biobank Axiom | Illumina OmniExpressExome Array |
| **Pre-imputation sample filters** | Call rate ≥95% | Call rate ≥97% | Call rate ≥95% | Call rate ≥95% | Call rate ≥95% | Call rate ≥99% |
| **Pre-imputation variant filters** | Call rate ≥95%<br>HWE *p*-value $\geq 1\times10^{-4}$<br>MAF ≥0.01 | Call rate ≥95%<br>HWE *p*-value $>1\times10^{-6}$<br>MAF >0.01 | Call rate ≥97%<br>HWE *p*-value $>1\times10^{-6}$<br>MAF >0.01 | Call rate ≥97%<br>HWE *p*-value $>1\times10^{-6}$<br>MAF >0.01 | Call rate ≥90%<br>HWE *p*-value $>1\times10^{-12}$<br>MAF >0.0001 | Call rate ≥95% |
| **Imputation panel** | Estonian Reference Panel | TOPMed | HRC | HRC | HRC and UK10K + 1000 Genomes phase 3 | 1000 Genomes phase 1 (version 3) |
| **Post-imputation variant filters** | Imputation quality ≥0.3; MAF ≥1%; MAC ≥20 | | | | | Imputation quality ≥0.3; MAF ≥1%; MAC ≥20; AF difference <0.2 compared to 1000 Genomes (Europeans) |
| **Association testing** | | | | | | |
| **Software** | SAIGE v0.43.1 | PLINK 2.0 | PLINK 2.0 | SNPTEST v2.5.4-beta3 | PLINK 2.0 | ProbABEL |
| **Model** | Logistic mixed model | Firth-fallback logistic regression | Firth-fallback logistic regression | Frequentist | Firth-fallback logistic regression | Logistic regression |
| **Related individuals** | Included | Excluded | Excluded | Excluded | Excluded | Excluded |
| **Covariates** | Age, age², sex, PCs 1-10 | Age, age², sex, PCs 1-10 | Age, age², sex, PCs 1-5 | Age, age², sex, PCs 1-10 | Age, age², sex, genotyping array, PCs 1-10 | Age, sex, source protocol, PCs 1-4 |

[*]Varenicline effectiveness phenotypes were defined using a combination of primary care EHRs (to obtain varenicline prescription events) and baseline questionnaire data (to obtain smoking status). [†]In GoDARTS, association testing was performed in individuals genotyped on each of the three different separately, and subsequently meta-analysed. [‡]Further description of the sample-level and variant-level genotyping quality control performed by individual cohort studies are available elsewhere for Estonian Biobank (Laisk *et al*[6]), EXCEED (Williams *et al*[7]), GoDARTS and GoSHARE (Srinivasan *et al*[8]) and UK Biobank (Bycroft *et al*[5]). *Abbreviations:* AF, allele frequency; EHR, electronic health record; HRC, Haplotype Reference Consortium; HWE, Hardy-Weinberg equilibrium; MAC, minor allele count; MAF, minor allele frequency; PC, principal component; TOPMed, Trans-Omics for Precision Medicine.

**Supplementary Table 3** Sentinel variants associated with **(A)** SVE and **(B)** LVE in Stage 1 GWASs.

| Variant | Chr:Position (hg19) | Info score* | Direction† | Heterogeneity estimate, $I^2$ (%) |
|---|---|---|---|---|
| **(A) Short-term varenicline effectiveness (SVE)** | | | | |
| rs186444103 | 2:40054022 | 0.982 | ???++ | 0 |
| rs4364036 | 2:162904624 | 0.900 | ????+ | 0 |
| rs4686373 | 3:9853437 | 0.998 | ?+?-+ | 49.9 |
| rs78597169 | 8:117830288 | 0.891 | ???-- | 0 |
| rs2297038 | 10:30663464 | 0.817 | ?+??+ | 0 |
| rs9600669 | 13:76776621 | 0.987 | ?-+++ | 43.0 |
| rs8003262 | 14:102050501 | 0.980 | ???++ | 0 |
| rs149318872 | 15:101162067 | 0.753 | ????- | 0 |
| rs10421326 | 19:473572 | 0.861 | ????+ | 0 |
| rs827937 | 20:46793101 | 0.991 | ?+??+ | 0 |
| **(B) Long-term varenicline effectiveness (LVE)** | | | | |
| rs895545 | 2:121721467 | 1.000 | ???-- | 0 |
| rs7712227 | 5:118656769 | 0.978 | ?+?-- | 0 |
| rs78695099 | 6:100195347 | 0.927 | ????- | 0 |
| rs35443123 | 7:85173800 | 0.990 | --??- | 8.2 |
| rs766169 | 22:47680100 | 1.000 | ???++ | 0 |

*Info score calculated in UK Biobank. †Direction column has the following order: Estonian Biobank, EXCEED, GoDARTS, GoSHARE, UK Biobank. *Abbreviations:* Chr, chromosome.

**Supplementary Table 4** 95% credible sets generated by fine-mapping of loci associated with **(A)** SVE and **(B)** LVE in Stage 1.

| Locus sentinel | Chr:Position (hg19) | Variant ID | Effect allele | Other allele | MAF | *N* | OR [95% CI] | *p*-value | PIP |
|---|---|---|---|---|---|---|---|---|---|
| **(A) Short-term varenicline effectiveness (SVE)** | | | | | | | | | |
| rs186444103 | 2:40054022 | rs186444103 | T | G | 0.026 | 3023 | 2.27 [1.61, 3.21] | 3.20E-06 | 0.3347 |
| rs186444103 | 2:40027750 | rs74900673 | T | C | 0.025 | 3023 | 2.26 [1.60, 3.18] | 3.74E-06 | 0.3113 |
| rs186444103 | 2:40062149 | rs115068901 | T | C | 0.025 | 3023 | 2.27 [1.61, 3.22] | 3.67E-06 | 0.3001 |
| rs186444103 | 2:40126792 | rs114238370 | T | C | 0.022 | 3023 | 2.18 [1.49, 3.18] | 5.37E-05 | 0.0539 |
| rs4364036 | 2:162904624 | rs4364036 | T | C | 0.034 | 2699 | 2.13 [1.55, 2.92] | 2.55E-06 | 0.6548 |
| rs4364036 | 2:162869850 | rs142456029 | A | G | 0.023 | 2699 | 2.47 [1.69, 3.62] | 3.28E-06 | 0.2289 |
| rs4364036 | 2:162818274 | rs79187915 | A | G | 0.030 | 2699 | 1.98 [1.42, 2.76] | 5.55E-05 | 0.0918 |
| rs4686373 | 3:9853437 | rs4686373 | A | G | 0.090 | 3167 | 1.55 [1.29, 1.85] | 2.50E-06 | 0.5511 |
| rs4686373 | 3:9809082 | rs293796 | T | C | 0.090 | 3167 | 1.49 [1.25, 1.79] | 1.20E-05 | 0.1642 |
| rs4686373 | 3:9852059 | rs34761997 | CGAT | C | 0.110 | 2699 | 0.71 [0.59, 0.84] | 1.11E-04 | 0.0294 |
| rs4686373 | 3:9842742 | rs6786980 | A | G | 0.112 | 3375 | 0.73 [0.62, 0.86] | 1.49E-04 | 0.0255 |
| rs4686373 | 3:9865173 | 3:9865173:G:GT | G | GT | 0.109 | 2699 | 0.71 [0.59, 0.85] | 1.33E-04 | 0.0254 |
| rs4686373 | 3:9867093 | rs6443268 | C | G | 0.111 | 3375 | 1.36 [1.15, 1.60] | 2.27E-04 | 0.0181 |
| rs4686373 | 3:9856892 | rs9836708 | T | C | 0.111 | 3375 | 1.36 [1.15, 1.60] | 2.45E-04 | 0.0171 |
| rs4686373 | 3:9858891 | rs58647240 | C | G | 0.111 | 3375 | 1.36 [1.15, 1.60] | 2.49E-04 | 0.0168 |
| rs4686373 | 3:9864641 | rs6766418 | A | C | 0.111 | 3375 | 1.36 [1.15, 1.60] | 2.52E-04 | 0.0167 |
| rs4686373 | 3:9864272 | rs7623315 | A | G | 0.111 | 3375 | 0.74 [0.63, 0.87] | 2.54E-04 | 0.0166 |
| rs4686373 | 3:9857371 | rs7610410 | A | T | 0.111 | 3375 | 0.74 [0.63, 0.87] | 2.56E-04 | 0.0165 |
| rs4686373 | 3:9857514 | rs7652460 | A | G | 0.111 | 3375 | 1.36 [1.15, 1.60] | 2.56E-04 | 0.0165 |
| rs4686373 | 3:9866064 | rs4686376 | C | G | 0.111 | 3375 | 1.36 [1.15, 1.60] | 2.58E-04 | 0.0162 |
| rs4686373 | 3:9861053 | rs3806666 | T | C | 0.111 | 3375 | 1.35 [1.15, 1.59] | 2.70E-04 | 0.0158 |
| rs4686373 | 3:10513705 | rs12637358 | C | G | 0.312 | 2765 | 0.80 [0.71, 0.91] | 3.25E-04 | 0.0156 |
| rs78597169 | 8:117830288 | rs78597169 | A | T | 0.026 | 3023 | 0.43 [0.30, 0.61] | 3.12E-06 | 0.5825 |
| rs78597169 | 8:118358227 | rs753006634 | A | AT | 0.382 | 2699 | 0.8 [0.70, 0.90] | 4.34E-04 | 0.1771 |
| rs78597169 | 8:117841402 | rs56777536 | T | C | 0.196 | 3441 | 1.24 [1.09, 1.40] | 9.89E-04 | 0.0887 |
| rs78597169 | 8:117841774 | rs10108200 | T | C | 0.196 | 3441 | 0.81 [0.71, 0.92] | 9.91E-04 | 0.0883 |
| rs78597169 | 8:117825404 | rs188539646 | A | G | 0.016 | 2699 | 2.23 [1.41, 3.52] | 5.68E-04 | 0.0202 |
| rs2297038 | 10:30663464 | rs2297038 | T | C | 0.434 | 2843 | 1.33 [1.18, 1.50] | 2.95E-06 | 0.9945 |
| rs9600669 | 13:76758048 | rs2175075 | A | G | 0.090 | 3375 | 0.64 [0.54, 0.77] | 2.00E-06 | 0.0487 |
| rs9600669 | 13:76776621 | rs9600669 | A | G | 0.086 | 3375 | 1.57 [1.30, 1.88] | 1.94E-06 | 0.0470 |
| rs9600669 | 13:76773963 | rs2328973 | C | G | 0.086 | 3375 | 0.64 [0.53, 0.77] | 1.95E-06 | 0.0467 |
| rs9600669 | 13:76766603 | rs6562946 | T | C | 0.090 | 3375 | 1.55 [1.29, 1.86] | 2.19E-06 | 0.0442 |
| rs9600669 | 13:76773368 | rs7983570 | C | G | 0.090 | 3375 | 0.65 [0.54, 0.77] | 2.24E-06 | 0.0438 |

| | | | | | | | | | |
|---|---|---|---|---|---|---|---|---|---|
| rs9600669 | 13:76759200 | rs56411210 | C | G | 0.086 | 3375 | 0.64 [0.53, 0.77] | 2.46E-06 | 0.0396 |
| rs9600669 | 13:76760147 | rs10507848 | A | G | 0.086 | 3375 | 1.56 [1.29, 1.87] | 2.48E-06 | 0.0394 |
| rs9600669 | 13:76781463 | rs9318413 | A | T | 0.091 | 3375 | 0.64 [0.54, 0.77] | 2.55E-06 | 0.0391 |
| rs9600669 | 13:76762700 | rs10507849 | T | C | 0.086 | 3375 | 0.64 [0.53, 0.77] | 2.52E-06 | 0.0386 |
| rs9600669 | 13:76753631 | rs12428519 | T | G | 0.090 | 3375 | 0.65 [0.54, 0.78] | 2.73E-06 | 0.0380 |
| rs9600669 | 13:76756563 | rs55918529 | T | C | 0.090 | 3375 | 0.65 [0.54, 0.78] | 2.71E-06 | 0.0376 |
| rs9600669 | 13:76755497 | rs73226165 | T | G | 0.090 | 3375 | 0.65 [0.54, 0.78] | 2.73E-06 | 0.0373 |
| rs9600669 | 13:76757485 | rs12427641 | A | G | 0.090 | 3375 | 1.54 [1.29, 1.85] | 2.75E-06 | 0.0373 |
| rs9600669 | 13:76748017 | rs2136340 | A | G | 0.087 | 3375 | 1.55 [1.29, 1.87] | 2.60E-06 | 0.0372 |
| rs9600669 | 13:76749009 | rs73226155 | T | C | 0.090 | 3375 | 1.54 [1.29, 1.85] | 2.81E-06 | 0.0368 |
| rs9600669 | 13:76749117 | rs73226156 | T | C | 0.090 | 3375 | 1.54 [1.29, 1.85] | 2.87E-06 | 0.0361 |
| rs9600669 | 13:76747030 | rs2036717 | T | C | 0.090 | 3375 | 1.54 [1.29, 1.85] | 2.91E-06 | 0.0356 |
| rs9600669 | 13:76748063 | rs2136339 | T | C | 0.090 | 3375 | 0.65 [0.54, 0.78] | 2.92E-06 | 0.0356 |
| rs9600669 | 13:76752411 | rs73226162 | T | C | 0.090 | 3375 | 1.54 [1.28, 1.84] | 2.97E-06 | 0.0354 |
| rs9600669 | 13:76746556 | rs11618016 | T | G | 0.090 | 3375 | 1.54 [1.28, 1.85] | 2.98E-06 | 0.0352 |
| rs9600669 | 13:76746743 | rs2036718 | T | C | 0.090 | 3375 | 1.54 [1.28, 1.85] | 2.96E-06 | 0.0352 |
| rs9600669 | 13:76744456 | rs59938144 | A | C | 0.090 | 3375 | 1.54 [1.28, 1.85] | 3.01E-06 | 0.0350 |
| rs9600669 | 13:76761187 | rs4142522 | A | G | 0.085 | 3375 | 1.56 [1.29, 1.87] | 3.07E-06 | 0.0323 |
| rs9600669 | 13:76753228 | rs12430486 | A | G | 0.090 | 3375 | 1.53 [1.28, 1.83] | 4.12E-06 | 0.0272 |
| rs9600669 | 13:76760227 | rs4142523 | T | C | 0.088 | 3375 | 0.65 [0.54, 0.78] | 4.14E-06 | 0.0267 |
| rs9600669 | 13:76741023 | rs534739393 | CA | C | 0.101 | 2843 | 0.67 [0.56, 0.81] | 2.38E-05 | 0.0067 |
| rs8003262 | 14:102050501 | rs8003262 | A | G | 0.059 | 3023 | 1.73 [1.37, 2.19] | 4.42E-06 | 0.2693 |
| rs8003262 | 14:102049001 | rs58241177 | A | C | 0.059 | 3023 | 0.58 [0.46, 0.73] | 4.51E-06 | 0.2640 |
| rs8003262 | 14:102046489 | rs11849688 | T | C | 0.059 | 3023 | 0.59 [0.46, 0.74] | 7.68E-06 | 0.1817 |
| rs8003262 | 14:102035570 | rs7158280 | T | C | 0.062 | 3023 | 1.60 [1.27, 2.00] | 5.50E-05 | 0.0491 |
| rs8003262 | 14:102034054 | rs10134976 | T | C | 0.062 | 3023 | 1.60 [1.27, 2.01] | 5.47E-05 | 0.0489 |
| rs8003262 | 14:102033210 | rs9324041 | A | G | 0.062 | 3023 | 0.63 [0.50, 0.79] | 6.11E-05 | 0.0451 |
| rs8003262 | 14:102033207 | rs9324040 | C | G | 0.061 | 3023 | 0.63 [0.50, 0.79] | 6.65E-05 | 0.0422 |
| rs8003262 | 14:102030527 | rs1997907 | C | G | 0.059 | 3023 | 0.62 [0.49, 0.78] | 6.22E-05 | 0.0415 |
| rs8003262 | 14:102048013 | rs33927302 | A | AT | 0.060 | 2699 | 1.66 [1.30, 2.12] | 5.56E-05 | 0.0402 |
| rs149318872 | 15:101162067 | rs149318872 | G | GGA | 0.049 | 2699 | 0.50 [0.37, 0.67] | 3.42E-06 | 0.5238 |
| rs149318872 | 15:101162066 | rs150515319 | T | TG | 0.094 | 2699 | 0.67 [0.55, 0.81] | 3.24E-05 | 0.3994 |
| rs149318872 | 15:101159421 | rs60819954 | T | C | 0.094 | 3265 | 0.73 [0.61, 0.88] | 6.09E-04 | 0.0462 |
| rs10421326 | 19:473572 | rs10421326 | C | G | 0.174 | 2699 | 1.53 [1.31, 1.78] | 5.74E-08 | 0.8160 |
| rs10421326 | 19:469979 | rs34770712 | A | AG | 0.189 | 2765 | 1.39 [1.21, 1.60] | 3.85E-06 | 0.0273 |
| rs10421326 | 19:470373 | 19:470373:A:AT | A | AT | 0.167 | 2699 | 0.70 [0.60, 0.82] | 4.08E-06 | 0.0235 |
| rs10421326 | 19:465427 | rs140502943 | G | GTGAA | 0.190 | 2765 | 1.37 [1.20, 1.57] | 5.21E-06 | 0.0221 |

| | | | | | | | | | |
|---|---|---|---|---|---|---|---|---|---|
| rs10421326 | 19:465302 | rs10406389 | A | G | 0.187 | 2699 | 0.73 [0.64, 0.84] | 6.56E-06 | 0.0176 |
| rs10421326 | 19:463326 | rs1968130 | A | G | 0.183 | 2909 | 1.36 [1.19, 1.56] | 9.35E-06 | 0.0134 |
| rs10421326 | 19:465071 | rs10405754 | A | G | 0.187 | 2843 | 0.74 [0.65, 0.85] | 1.12E-05 | 0.0113 |
| rs10421326 | 19:466793 | rs10414788 | A | G | 0.186 | 2843 | 0.74 [0.64, 0.84] | 1.16E-05 | 0.0110 |
| rs10421326 | 19:465889 | rs28972633 | T | C | 0.216 | 2699 | 0.75 [0.66, 0.85] | 1.33E-05 | 0.0101 |
| rs827937 | 20:46793101 | rs827937 | A | T | 0.248 | 2843 | 1.37 [1.21, 1.55] | 1.08E-06 | 0.2062 |
| rs827937 | 20:46792921 | rs2605205 | A | G | 0.247 | 2843 | 1.37 [1.20, 1.55] | 1.29E-06 | 0.1761 |
| rs827937 | 20:46788676 | rs228194 | A | G | 0.245 | 3441 | 1.33 [1.18, 1.50] | 2.33E-06 | 0.1093 |
| rs827937 | 20:46787825 | rs827944 | T | C | 0.244 | 3441 | 0.75 [0.67, 0.85] | 2.46E-06 | 0.1033 |
| rs827937 | 20:46787378 | rs827947 | C | G | 0.244 | 3441 | 1.33 [1.18, 1.49] | 2.80E-06 | 0.0941 |
| rs827937 | 20:46787459 | rs827946 | T | G | 0.245 | 3441 | 0.76 [0.67, 0.85] | 3.11E-06 | 0.0863 |
| rs827937 | 20:46792944 | rs827939 | T | C | 0.245 | 3375 | 0.76 [0.67, 0.85] | 4.61E-06 | 0.0596 |
| rs827937 | 20:46792236 | rs827941 | T | C | 0.245 | 3441 | 0.76 [0.67, 0.85] | 4.78E-06 | 0.0586 |
| rs827937 | 20:46793073 | rs67232918 | A | ACACT | 0.250 | 2699 | 0.74 [0.65, 0.84] | 5.30E-06 | 0.0500 |
| rs827937 | 20:46789064 | 20:46789064:A:AAAACAAACAAAC | A | AAAACAAACAAAC | 0.188 | 2699 | 1.38 [1.20, 1.59] | 8.63E-06 | 0.0299 |
| **(B) Long-term varenicline effectiveness (LVE)** | | | | | | | | | |
| rs895545 | 2:121721467 | rs895545 | T | C | 0.057 | 3513 | 0.59 [0.48, 0.73] | 1.21E-06 | 0.0487 |
| rs895545 | 2:121720125 | rs4848664 | T | C | 0.057 | 3513 | 1.69 [1.36, 2.09] | 1.64E-06 | 0.0388 |
| rs895545 | 2:121721845 | rs895546 | A | T | 0.056 | 3687 | 0.60 [0.48, 0.74] | 1.83E-06 | 0.0364 |
| rs895545 | 2:121723791 | rs11122838 | T | C | 0.057 | 3687 | 0.60 [0.49, 0.74] | 2.07E-06 | 0.0350 |
| rs895545 | 2:121719268 | rs13432147 | T | C | 0.057 | 3687 | 1.66 [1.34, 2.05] | 2.50E-06 | 0.0299 |
| rs895545 | 2:121725452 | rs2677527 | A | G | 0.057 | 3687 | 0.61 [0.49, 0.75] | 2.79E-06 | 0.0281 |
| rs895545 | 2:121725534 | rs2592596 | T | C | 0.057 | 3687 | 0.61 [0.49, 0.75] | 2.78E-06 | 0.0281 |
| rs895545 | 2:121726927 | rs2677509 | T | C | 0.057 | 3687 | 0.61 [0.49, 0.75] | 2.79E-06 | 0.0281 |
| rs895545 | 2:121725418 | rs2677528 | A | T | 0.057 | 3687 | 1.65 [1.34, 2.03] | 2.80E-06 | 0.0280 |
| rs895545 | 2:121719538 | rs2166562 | A | G | 0.060 | 3336 | 0.60 [0.49, 0.74] | 2.71E-06 | 0.0274 |
| rs895545 | 2:121719537 | rs2166563 | A | C | 0.060 | 3336 | 1.66 [1.34, 2.06] | 2.79E-06 | 0.0268 |
| rs895545 | 2:121718926 | rs4422162 | A | G | 0.056 | 3687 | 1.66 [1.34, 2.06] | 2.94E-06 | 0.0255 |
| rs895545 | 2:121729323 | rs2677507 | A | G | 0.057 | 3687 | 1.64 [1.33, 2.03] | 3.68E-06 | 0.0224 |
| rs895545 | 2:121719089 | rs4625910 | A | G | 0.057 | 3687 | 0.61 [0.50, 0.75] | 3.93E-06 | 0.0219 |
| rs895545 | 2:121720245 | rs4848667 | T | C | 0.057 | 3687 | 1.64 [1.33, 2.02] | 4.03E-06 | 0.0212 |
| rs895545 | 2:121718978 | rs4287773 | T | C | 0.057 | 3687 | 1.64 [1.33, 2.02] | 4.11E-06 | 0.0209 |
| rs895545 | 2:121719171 | rs4625911 | A | G | 0.057 | 3687 | 0.61 [0.50, 0.75] | 4.12E-06 | 0.0208 |
| rs895545 | 2:121719195 | rs4625912 | A | G | 0.057 | 3687 | 0.61 [0.50, 0.75] | 4.11E-06 | 0.0208 |
| rs895545 | 2:121719948 | rs2121435 | T | C | 0.057 | 3687 | 1.64 [1.33, 2.02] | 4.12E-06 | 0.0208 |
| rs895545 | 2:121720126 | rs4848665 | A | G | 0.057 | 3687 | 0.61 [0.50, 0.75] | 4.12E-06 | 0.0208 |
| rs895545 | 2:121720191 | rs4848666 | T | C | 0.057 | 3687 | 0.61 [0.50, 0.75] | 4.12E-06 | 0.0208 |

| | | | | | | | | | |
|---|---|---|---|---|---|---|---|---|---|
| rs895545 | 2:121720300 | rs4848668 | T | C | 0.057 | 3687 | 0.61 [0.50, 0.75] | 4.12E-06 | 0.0208 |
| rs895545 | 2:121720392 | rs4848669 | T | C | 0.057 | 3687 | 1.64 [1.33, 2.02] | 4.12E-06 | 0.0208 |
| rs895545 | 2:121720620 | rs7349266 | A | G | 0.057 | 3687 | 1.64 [1.33, 2.02] | 4.12E-06 | 0.0208 |
| rs895545 | 2:121709536 | rs12711535 | T | C | 0.057 | 3687 | 0.61 [0.49, 0.75] | 4.13E-06 | 0.0203 |
| rs895545 | 2:121710855 | rs2084234 | T | G | 0.057 | 3687 | 1.64 [1.33, 2.02] | 4.44E-06 | 0.0197 |
| rs895545 | 2:121713030 | rs12614482 | A | G | 0.057 | 3687 | 0.61 [0.50, 0.76] | 4.70E-06 | 0.0188 |
| rs895545 | 2:121720434 | rs11680138 | A | T | 0.057 | 3687 | 1.63 [1.32, 2.01] | 4.71E-06 | 0.0188 |
| rs895545 | 2:121713203 | rs13010895 | A | G | 0.057 | 3687 | 0.61 [0.50, 0.76] | 4.71E-06 | 0.0188 |
| rs895545 | 2:121729172 | rs2243855 | T | C | 0.057 | 3687 | 0.61 [0.50, 0.76] | 4.85E-06 | 0.0185 |
| rs895545 | 2:121709786 | rs12711536 | T | C | 0.057 | 3687 | 0.61 [0.50, 0.76] | 4.86E-06 | 0.0184 |
| rs895545 | 2:121716605 | rs753900 | C | G | 0.057 | 3687 | 1.63 [1.32, 2.02] | 4.87E-06 | 0.0183 |
| rs895545 | 2:121712361 | rs4848660 | T | C | 0.057 | 3687 | 1.63 [1.32, 2.01] | 4.92E-06 | 0.0181 |
| rs895545 | 2:121712381 | rs4848661 | T | C | 0.057 | 3687 | 0.61 [0.50, 0.76] | 4.92E-06 | 0.0181 |
| rs895545 | 2:121712418 | rs4848662 | A | G | 0.057 | 3687 | 0.61 [0.50, 0.76] | 4.91E-06 | 0.0181 |
| rs895545 | 2:121712441 | rs4848663 | T | C | 0.057 | 3687 | 1.63 [1.32, 2.01] | 4.92E-06 | 0.0181 |
| rs895545 | 2:121712507 | rs4848134 | A | C | 0.057 | 3687 | 1.63 [1.32, 2.01] | 4.92E-06 | 0.0181 |
| rs895545 | 2:121711860 | rs11122837 | A | G | 0.057 | 3687 | 0.61 [0.50, 0.76] | 4.94E-06 | 0.0181 |
| rs895545 | 2:121712260 | rs11689760 | A | C | 0.057 | 3687 | 1.63 [1.32, 2.01] | 4.94E-06 | 0.0181 |
| rs895545 | 2:121713041 | rs12617878 | T | C | 0.057 | 3687 | 1.63 [1.32, 2.01] | 5.19E-06 | 0.0176 |
| rs895545 | 2:121719392 | rs7420788 | A | G | 0.058 | 3687 | 1.59 [1.29, 1.95] | 1.19E-05 | 0.0098 |
| rs895545 | 2:121692477 | rs2166565 | A | G | 0.052 | 3687 | 1.58 [1.27, 1.97] | 4.53E-05 | 0.0032 |
| rs895545 | 2:121695699 | rs895550 | A | G | 0.054 | 3687 | 0.64 [0.52, 0.79] | 4.98E-05 | 0.0031 |
| rs895545 | 2:121708159 | rs11122835 | T | G | 0.053 | 3687 | 1.57 [1.26, 1.96] | 4.97E-05 | 0.0030 |
| rs895545 | 2:121709091 | rs11681136 | A | G | 0.053 | 3687 | 0.64 [0.51, 0.79] | 5.03E-05 | 0.0030 |
| rs7712227 | 5:118656769 | rs7712227 | A | G | 0.060 | 3687 | 0.59 [0.47, 0.73] | 2.56E-06 | 0.0317 |
| rs7712227 | 5:118671118 | rs7725244 | A | G | 0.062 | 3886 | 0.61 [0.50, 0.75] | 4.33E-06 | 0.0248 |
| rs7712227 | 5:118666751 | rs72786143 | T | C | 0.062 | 3886 | 1.63 [1.32, 2.02] | 4.80E-06 | 0.0231 |
| rs7712227 | 5:118670540 | rs72786161 | T | G | 0.062 | 3886 | 1.63 [1.32, 2.02] | 4.75E-06 | 0.0230 |
| rs7712227 | 5:118659359 | rs72786136 | A | G | 0.061 | 3886 | 0.62 [0.50, 0.76] | 7.42E-06 | 0.0167 |
| rs7712227 | 5:118675017 | rs7737934 | T | C | 0.063 | 3886 | 1.60 [1.30, 1.97] | 8.09E-06 | 0.0162 |
| rs7712227 | 5:118660784 | rs66535134 | A | T | 0.062 | 3687 | 1.62 [1.31, 2.01] | 7.88E-06 | 0.0156 |
| rs7712227 | 5:118675446 | rs3813305 | T | G | 0.050 | 3513 | 1.76 [1.38, 2.24] | 4.89E-06 | 0.0145 |
| rs7712227 | 5:118667642 | rs7704398 | T | C | 0.063 | 3886 | 0.63 [0.51, 0.77] | 1.04E-05 | 0.0136 |
| rs7712227 | 5:118674701 | rs7724178 | T | C | 0.063 | 3886 | 1.59 [1.30, 1.96] | 1.06E-05 | 0.0133 |
| rs7712227 | 5:118674893 | rs7704334 | A | G | 0.063 | 3886 | 1.59 [1.30, 1.96] | 1.06E-05 | 0.0133 |
| rs7712227 | 5:118674923 | rs7704487 | T | C | 0.063 | 3886 | 0.63 [0.51, 0.77] | 1.06E-05 | 0.0133 |
| rs7712227 | 5:118674774 | rs7724204 | T | C | 0.063 | 3886 | 1.59 [1.30, 1.96] | 1.06E-05 | 0.0133 |

| | | | | | | | | | |
|---|---|---|---|---|---|---|---|---|---|
| rs7712227 | 5:118674623 | rs7723976 | T | C | 0.063 | 3886 | 1.59 [1.30, 1.96] | 1.07E-05 | 0.0133 |
| rs7712227 | 5:118674652 | rs7704291 | A | G | 0.063 | 3886 | 0.63 [0.51, 0.77] | 1.06E-05 | 0.0133 |
| rs7712227 | 5:118674284 | rs6862831 | C | G | 0.063 | 3886 | 0.63 [0.51, 0.77] | 1.12E-05 | 0.0129 |
| rs7712227 | 5:118674290 | rs6862517 | C | G | 0.063 | 3886 | 1.59 [1.29, 1.96] | 1.12E-05 | 0.0128 |
| rs7712227 | 5:118673697 | rs17164572 | T | C | 0.063 | 3886 | 0.63 [0.51, 0.77] | 1.14E-05 | 0.0126 |
| rs7712227 | 5:118672441 | rs17145198 | A | G | 0.063 | 3886 | 1.59 [1.29, 1.96] | 1.15E-05 | 0.0125 |
| rs7712227 | 5:118672472 | rs11955852 | A | C | 0.063 | 3886 | 1.59 [1.29, 1.96] | 1.15E-05 | 0.0125 |
| rs7712227 | 5:118661434 | rs111332768 | T | C | 0.064 | 3886 | 1.58 [1.29, 1.94] | 1.19E-05 | 0.0125 |
| rs7712227 | 5:118661649 | rs67311526 | T | C | 0.064 | 3886 | 1.58 [1.29, 1.95] | 1.18E-05 | 0.0124 |
| rs7712227 | 5:118674055 | rs6862320 | A | G | 0.063 | 3886 | 0.63 [0.51, 0.77] | 1.16E-05 | 0.0124 |
| rs7712227 | 5:118674747 | rs757741772 | CA | C | 0.061 | 3162 | 0.58 [0.46, 0.74] | 6.96E-06 | 0.0123 |
| rs7712227 | 5:118669224 | rs11959659 | A | G | 0.063 | 3886 | 0.63 [0.51, 0.77] | 1.19E-05 | 0.0123 |
| rs7712227 | 5:118671834 | rs60507834 | A | G | 0.063 | 3886 | 1.59 [1.29, 1.95] | 1.20E-05 | 0.0123 |
| rs7712227 | 5:118672058 | rs17145194 | A | G | 0.063 | 3886 | 1.59 [1.29, 1.95] | 1.20E-05 | 0.0123 |
| rs7712227 | 5:118672137 | rs17145196 | A | C | 0.063 | 3886 | 0.63 [0.51, 0.77] | 1.20E-05 | 0.0123 |
| rs7712227 | 5:118674041 | rs6883394 | T | C | 0.063 | 3886 | 1.59 [1.29, 1.96] | 1.18E-05 | 0.0122 |
| rs7712227 | 5:118669780 | rs72786157 | A | G | 0.063 | 3886 | 1.59 [1.29, 1.95] | 1.21E-05 | 0.0122 |
| rs7712227 | 5:118671110 | rs7707221 | T | C | 0.063 | 3886 | 1.59 [1.29, 1.95] | 1.21E-05 | 0.0122 |
| rs7712227 | 5:118668406 | rs6868702 | A | G | 0.063 | 3886 | 1.59 [1.29, 1.95] | 1.21E-05 | 0.0121 |
| rs7712227 | 5:118668538 | rs6869017 | C | G | 0.063 | 3886 | 1.59 [1.29, 1.95] | 1.21E-05 | 0.0121 |
| rs7712227 | 5:118672197 | rs73790807 | A | G | 0.063 | 3886 | 1.59 [1.29, 1.95] | 1.21E-05 | 0.0121 |
| rs7712227 | 5:118668381 | rs6868688 | A | C | 0.063 | 3886 | 1.59 [1.29, 1.95] | 1.21E-05 | 0.0121 |
| rs7712227 | 5:118670591 | rs17145189 | A | G | 0.063 | 3886 | 0.63 [0.51, 0.77] | 1.21E-05 | 0.0121 |
| rs7712227 | 5:118671062 | rs11951771 | T | G | 0.063 | 3886 | 1.59 [1.29, 1.95] | 1.21E-05 | 0.0121 |
| rs7712227 | 5:118667909 | rs72786149 | T | G | 0.063 | 3886 | 1.59 [1.29, 1.95] | 1.21E-05 | 0.0121 |
| rs7712227 | 5:118667979 | rs72786152 | C | G | 0.063 | 3886 | 1.59 [1.29, 1.95] | 1.21E-05 | 0.0121 |
| rs7712227 | 5:118671462 | rs7707736 | T | C | 0.063 | 3886 | 1.59 [1.29, 1.95] | 1.20E-05 | 0.0120 |
| rs7712227 | 5:118671517 | rs7725878 | A | G | 0.063 | 3886 | 0.63 [0.51, 0.77] | 1.20E-05 | 0.0120 |
| rs7712227 | 5:118667610 | rs7704386 | C | G | 0.063 | 3886 | 1.59 [1.29, 1.95] | 1.21E-05 | 0.0120 |
| rs7712227 | 5:118667072 | rs72786144 | A | T | 0.063 | 3886 | 0.63 [0.51, 0.78] | 1.22E-05 | 0.0120 |
| rs7712227 | 5:118667211 | rs7703998 | A | G | 0.063 | 3886 | 0.63 [0.51, 0.78] | 1.22E-05 | 0.0120 |
| rs7712227 | 5:118666627 | rs72786142 | A | C | 0.063 | 3886 | 0.63 [0.51, 0.78] | 1.22E-05 | 0.0119 |
| rs7712227 | 5:118663397 | rs7711312 | A | G | 0.063 | 3886 | 0.63 [0.51, 0.78] | 1.23E-05 | 0.0119 |
| rs7712227 | 5:118674020 | rs6862299 | A | G | 0.063 | 3886 | 0.63 [0.51, 0.77] | 1.23E-05 | 0.0119 |
| rs7712227 | 5:118667253 | rs7704026 | A | G | 0.063 | 3886 | 0.63 [0.51, 0.78] | 1.24E-05 | 0.0119 |
| rs7712227 | 5:118674026 | rs6861849 | A | G | 0.063 | 3886 | 1.59 [1.29, 1.96] | 1.25E-05 | 0.0118 |
| rs7712227 | 5:118663310 | rs11958957 | T | C | 0.063 | 3886 | 0.63 [0.51, 0.78] | 1.25E-05 | 0.0118 |

| | | | | | | | | | |
|---|---|---|---|---|---|---|---|---|---|
| rs7712227 | 5:118663373 | rs7710939 | T | C | 0.063 | 3886 | 0.63 [0.51, 0.78] | 1.25E-05 | 0.0118 |
| rs7712227 | 5:118674429 | rs7703577 | A | C | 0.063 | 3886 | 1.59 [1.29, 1.95] | 1.26E-05 | 0.0115 |
| rs7712227 | 5:118675246 | rs3813301 | A | G | 0.063 | 3886 | 1.58 [1.29, 1.95] | 1.31E-05 | 0.0113 |
| rs7712227 | 5:118661206 | rs181617086 | A | G | 0.063 | 3886 | 0.64 [0.52, 0.78] | 1.79E-05 | 0.0091 |
| rs7712227 | 5:118667451 | rs7724082 | T | C | 0.064 | 3886 | 1.57 [1.28, 1.92] | 1.87E-05 | 0.0089 |
| rs7712227 | 5:118671091 | rs11954854 | A | C | 0.064 | 3886 | 1.57 [1.28, 1.93] | 1.88E-05 | 0.0088 |
| rs7712227 | 5:118660500 | rs11956285 | T | G | 0.063 | 3886 | 0.64 [0.52, 0.78] | 1.87E-05 | 0.0087 |
| rs7712227 | 5:118660569 | rs11959729 | T | C | 0.063 | 3886 | 1.57 [1.28, 1.93] | 1.87E-05 | 0.0087 |
| rs7712227 | 5:118659537 | rs60161062 | A | G | 0.063 | 3886 | 0.64 [0.52, 0.78] | 1.87E-05 | 0.0087 |
| rs7712227 | 5:119246341 | rs34865044 | A | AT | 0.247 | 3535 | 1.27 [1.13, 1.43] | 4.60E-05 | 0.0084 |
| rs7712227 | 5:118660289 | rs11956215 | A | G | 0.063 | 3886 | 0.64 [0.52, 0.78] | 2.01E-05 | 0.0082 |
| rs7712227 | 5:118658731 | rs7722699 | A | G | 0.062 | 3886 | 0.64 [0.52, 0.78] | 2.04E-05 | 0.0081 |
| rs7712227 | 5:118658702 | rs7722687 | T | G | 0.063 | 3886 | 0.64 [0.52, 0.78] | 2.09E-05 | 0.0080 |
| rs7712227 | 5:118637787 | rs72786120 | A | G | 0.039 | 3513 | 0.53 [0.40, 0.70] | 9.01E-06 | 0.0060 |
| rs7712227 | 5:118659296 | rs60756572 | C | G | 0.063 | 3886 | 1.55 [1.26, 1.90] | 3.25E-05 | 0.0058 |
| rs7712227 | 5:118628651 | rs60599743 | A | T | 0.054 | 3687 | 0.61 [0.48, 0.77] | 2.53E-05 | 0.0053 |
| rs7712227 | 5:118672213 | 5:118672213:G:GACTTCTTAT | G | GACTTCTTAT | 0.064 | 3535 | 0.63 [0.51, 0.79] | 3.65E-05 | 0.0048 |
| rs7712227 | 5:118675405 | rs3813304 | A | G | 0.051 | 3712 | 1.63 [1.29, 2.06] | 3.66E-05 | 0.0040 |
| rs7712227 | 5:118661697 | rs11960654 | T | C | 0.066 | 3886 | 1.51 [1.23, 1.84] | 6.06E-05 | 0.0039 |
| rs7712227 | 5:118665726 | rs67154545 | A | G | 0.066 | 3886 | 1.51 [1.23, 1.85] | 6.30E-05 | 0.0037 |
| rs7712227 | 5:118665801 | rs11956042 | C | G | 0.066 | 3886 | 1.51 [1.23, 1.85] | 6.30E-05 | 0.0037 |
| rs7712227 | 5:118666249 | rs11960246 | T | C | 0.066 | 3886 | 1.51 [1.23, 1.85] | 6.30E-05 | 0.0037 |
| rs7712227 | 5:118666603 | rs72786141 | A | G | 0.066 | 3886 | 1.51 [1.23, 1.85] | 6.26E-05 | 0.0037 |
| rs7712227 | 5:118665520 | rs11959451 | T | G | 0.066 | 3886 | 1.51 [1.23, 1.85] | 6.31E-05 | 0.0037 |
| rs7712227 | 5:118665158 | rs4392658 | T | C | 0.066 | 3886 | 1.51 [1.23, 1.85] | 6.34E-05 | 0.0037 |
| rs7712227 | 5:118665421 | rs1876737 | C | G | 0.066 | 3886 | 0.66 [0.54, 0.81] | 6.32E-05 | 0.0037 |
| rs7712227 | 5:118663542 | rs7711101 | A | C | 0.066 | 3886 | 1.51 [1.23, 1.85] | 6.33E-05 | 0.0037 |
| rs7712227 | 5:118663979 | rs6875361 | A | T | 0.066 | 3886 | 1.51 [1.23, 1.85] | 6.34E-05 | 0.0037 |
| rs7712227 | 5:118664573 | rs11950366 | T | C | 0.066 | 3886 | 1.51 [1.23, 1.85] | 6.34E-05 | 0.0037 |
| rs7712227 | 5:118664645 | rs11959964 | T | C | 0.066 | 3886 | 0.66 [0.54, 0.81] | 6.34E-05 | 0.0037 |
| rs7712227 | 5:118665062 | rs67708265 | A | G | 0.066 | 3886 | 1.51 [1.23, 1.85] | 6.34E-05 | 0.0037 |
| rs7712227 | 5:118670361 | rs72786159 | T | C | 0.064 | 3535 | 1.56 [1.26, 1.93] | 5.34E-05 | 0.0037 |
| rs7712227 | 5:118672208 | rs111815603 | C | G | 0.064 | 3535 | 1.56 [1.26, 1.93] | 5.34E-05 | 0.0037 |
| rs7712227 | 5:118670187 | 5:118670187:A:AC | A | AC | 0.064 | 3535 | 0.64 [0.52, 0.80] | 5.34E-05 | 0.0037 |
| rs7712227 | 5:118627151 | rs73794110 | T | C | 0.053 | 3886 | 0.63 [0.50, 0.79] | 5.44E-05 | 0.0033 |
| rs7712227 | 5:118649135 | rs7701688 | A | T | 0.052 | 3687 | 1.62 [1.28, 2.04] | 5.02E-05 | 0.0033 |
| rs7712227 | 5:118616559 | rs67186626 | T | C | 0.053 | 3886 | 0.63 [0.50, 0.79] | 5.91E-05 | 0.0031 |

| | | | | | | | | | |
|---|---|---|---|---|---|---|---|---|---|
| rs7712227 | 5:118621929 | rs73794107 | A | G | 0.053 | 3886 | 1.58 [1.27, 1.98] | 5.99E-05 | 0.0031 |
| rs7712227 | 5:118620885 | rs72786113 | T | C | 0.053 | 3886 | 1.58 [1.27, 1.98] | 6.05E-05 | 0.0031 |
| rs7712227 | 5:118628798 | rs72786118 | A | G | 0.053 | 3886 | 1.58 [1.26, 1.98] | 6.28E-05 | 0.0030 |
| rs7712227 | 5:118661203 | rs188811851 | T | C | 0.066 | 3886 | 1.50 [1.22, 1.83] | 8.78E-05 | 0.0029 |
| rs7712227 | 5:119291969 | rs34618848 | G | GA | 0.388 | 3535 | 1.22 [1.10, 1.34] | 1.64E-04 | 0.0028 |
| rs7712227 | 5:118619536 | rs10519576 | A | G | 0.053 | 3886 | 0.63 [0.51, 0.79] | 6.78E-05 | 0.0028 |
| rs7712227 | 5:118647014 | rs58131651 | T | C | 0.053 | 3886 | 0.63 [0.50, 0.79] | 6.75E-05 | 0.0028 |
| rs7712227 | 5:118635627 | rs6892643 | C | G | 0.053 | 3886 | 1.58 [1.26, 1.97] | 6.93E-05 | 0.0028 |
| rs7712227 | 5:118635662 | rs6892980 | A | G | 0.053 | 3886 | 0.63 [0.51, 0.79] | 6.93E-05 | 0.0028 |
| rs7712227 | 5:118640218 | rs6879962 | A | T | 0.053 | 3886 | 1.58 [1.26, 1.97] | 7.01E-05 | 0.0028 |
| rs7712227 | 5:118637058 | rs56319623 | A | G | 0.053 | 3886 | 0.63 [0.51, 0.79] | 7.00E-05 | 0.0028 |
| rs7712227 | 5:118643238 | rs72786122 | A | G | 0.053 | 3886 | 0.63 [0.50, 0.79] | 6.89E-05 | 0.0028 |
| rs7712227 | 5:118664034 | rs11421168 | CA | C | 0.066 | 3162 | 0.64 [0.51, 0.80] | 7.45E-05 | 0.0028 |
| rs7712227 | 5:118639499 | rs10519581 | A | G | 0.053 | 3886 | 1.58 [1.26, 1.97] | 7.03E-05 | 0.0028 |
| rs7712227 | 5:118646065 | rs72786125 | T | C | 0.053 | 3886 | 0.63 [0.50, 0.79] | 6.93E-05 | 0.0028 |
| rs7712227 | 5:118645320 | rs17145135 | A | G | 0.053 | 3886 | 0.63 [0.51, 0.79] | 7.14E-05 | 0.0027 |
| rs7712227 | 5:118626215 | rs72786116 | A | G | 0.053 | 3886 | 0.63 [0.50, 0.79] | 7.03E-05 | 0.0027 |
| rs7712227 | 5:118677967 | rs67967956 | C | G | 0.061 | 3886 | 1.52 [1.23, 1.88] | 9.58E-05 | 0.0025 |
| rs78695099 | 6:99997165 | rs2894870 | A | T | 0.036 | 3162 | 0.47 [0.35, 0.63] | 2.84E-07 | 0.3621 |
| rs78695099 | 6:99997166 | rs2894871 | A | C | 0.035 | 3162 | 0.48 [0.36, 0.64] | 5.09E-07 | 0.2473 |
| rs78695099 | 6:100195347 | rs78695099 | A | G | 0.026 | 3162 | 0.42 [0.30, 0.58] | 2.05E-07 | 0.2030 |
| rs78695099 | 6:100022491 | rs75713647 | T | C | 0.025 | 3162 | 2.21 [1.60, 3.07] | 1.81E-06 | 0.0623 |
| rs78695099 | 6:100271658 | rs540690087 | A | G | 0.074 | 3162 | 1.50 [1.23, 1.83] | 5.45E-05 | 0.0349 |
| rs78695099 | 6:99901767 | rs148375451 | T | C | 0.026 | 3513 | 2.02 [1.48, 2.76] | 9.90E-06 | 0.0285 |
| rs78695099 | 6:100200753 | rs74401013 | T | C | 0.088 | 3687 | 1.38 [1.17, 1.63] | 1.42E-04 | 0.0211 |
| rs35443123 | 7:85173800 | rs35443123 | A | AT | 0.434 | 3535 | 0.79 [0.72, 0.88] | 4.86E-06 | 0.6095 |
| rs35443123 | 7:85142693 | rs142645420 | CCTAT | C | 0.421 | 3361 | 0.81 [0.73, 0.90] | 6.04E-05 | 0.0640 |
| rs35443123 | 7:85157517 | rs9942699 | A | C | 0.388 | 4131 | 1.21 [1.10, 1.33] | 7.17E-05 | 0.0536 |
| rs35443123 | 7:85157662 | rs6953439 | A | G | 0.423 | 4131 | 1.20 [1.09, 1.32] | 1.47E-04 | 0.0285 |
| rs35443123 | 7:85166174 | rs35499102 | A | G | 0.423 | 4131 | 0.84 [0.76, 0.92] | 1.59E-04 | 0.0263 |
| rs35443123 | 7:85160755 | rs6951823 | T | G | 0.423 | 4131 | 1.20 [1.09, 1.31] | 1.66E-04 | 0.0259 |
| rs35443123 | 7:85174307 | rs28432363 | A | G | 0.431 | 4131 | 0.84 [0.76, 0.92] | 1.73E-04 | 0.0249 |
| rs35443123 | 7:85168495 | rs13230012 | T | C | 0.314 | 3535 | 0.82 [0.74, 0.91] | 2.18E-04 | 0.0204 |
| rs35443123 | 7:85004755 | rs77649879 | T | C | 0.235 | 4093 | 1.23 [1.10, 1.37] | 2.63E-04 | 0.0174 |
| rs35443123 | 7:85177938 | rs10252600 | T | C | 0.430 | 4131 | 1.19 [1.08, 1.30] | 3.56E-04 | 0.0130 |
| rs35443123 | 7:85116129 | rs12704136 | T | C | 0.426 | 4131 | 0.84 [0.77, 0.93] | 4.34E-04 | 0.0108 |
| rs35443123 | 7:85113311 | rs1554847 | A | G | 0.427 | 4131 | 1.18 [1.08, 1.30] | 4.75E-04 | 0.0102 |

| | | | | | | | | | |
|---|---|---|---|---|---|---|---|---|---|
| rs35443123 | 7:85123452 | rs35923742 | A | T | 0.426 | 4131 | 1.18 [1.08, 1.30] | 5.24E-04 | 0.0094 |
| rs35443123 | 7:85112099 | rs55662263 | T | C | 0.427 | 4131 | 0.85 [0.77, 0.93] | 5.53E-04 | 0.0089 |
| rs35443123 | 7:85110875 | rs55804990 | A | G | 0.427 | 4131 | 1.18 [1.07, 1.30] | 5.65E-04 | 0.0088 |
| rs35443123 | 7:85120974 | rs34596135 | T | C | 0.426 | 3932 | 1.18 [1.08, 1.30] | 5.71E-04 | 0.0087 |
| rs35443123 | 7:85121064 | rs10499889 | T | C | 0.427 | 4131 | 1.18 [1.07, 1.29] | 6.11E-04 | 0.0082 |
| rs35443123 | 7:85155661 | rs2140458 | C | G | 0.323 | 4131 | 1.19 [1.07, 1.31] | 6.47E-04 | 0.0077 |
| rs766169 | 22:47680100 | rs766169 | T | G | 0.051 | 3513 | 1.84 [1.47, 2.29] | 6.68E-08 | 0.6043 |
| rs766169 | 22:47679901 | rs766168 | T | C | 0.050 | 3513 | 1.79 [1.43, 2.23] | 2.70E-07 | 0.2100 |
| rs766169 | 22:47679506 | rs58656950 | A | G | 0.049 | 3513 | 1.80 [1.44, 2.25] | 3.12E-07 | 0.1809 |

*Abbreviations:* MAF, minor allele frequency; N, sample size; OR, odds ratio; PIP, posterior inclusion probability.

**Supplementary Table 5** Look-up of variants identified in association studies of varenicline-related phenotypes and smoking cessation in SVE and LVE Stage 1 GWASs.

| Reference | Variant | Chr:Position (hg19) | Gene | EA | Stage 1 GWAS of SVE | | | Stage 1 GWAS of LVE | | |
|---|---|---|---|---|---|---|---|---|---|---|
| | | | | | EAF | OR [95% CI] | *p*-value | EAF | OR [95% CI] | *p*-value |
| **Varenicline efficacy (from candidate gene studies)** | | | | | | | | | | |
| [9] | rs4292956 | 1:154548946 | *CHRNB2* | T | 0.075 | 0.97 [0.79, 1.18] | $7.59\times10^{-1}$ | 0.073 | 1.03 [0.86, 1.24] | $7.19\times10^{-1}$ |
| [9] | rs3811450 | 1:154551032 | *CHRNB2* | T | 0.076 | 0.96 [0.79, 1.17] | $6.73\times10^{-1}$ | 0.074 | 1.01 [0.84, 1.21] | $9.42\times10^{-1}$ |
| [9] | rs6494212 | 15:32385119 | *CHRNA7* | T | 0.317 | 0.93 [0.83, 1.04] | $2.10\times10^{-1}$ | 0.314 | 1.05 [0.95, 1.17] | $3.59\times10^{-1}$ |
| [9] | rs2938674 | 15:78757913 | *IREB2* | A | 0.205 | 1.00 [0.88, 1.13] | $9.95\times10^{-1}$ | 0.206 | 1.07 [0.96, 1.20] | $2.08\times10^{-1}$ |
| [9] | rs7164594 | 15:78803057 | *HYKK* | T | 0.205 | 1.04 [0.92, 1.18] | $5.09\times10^{-1}$ | 0.204 | 1.10 [0.98, 1.24] | $9.16\times10^{-2}$ |
| [10] | rs16969968 | 15:78882925 | *CHRNA5* | A | 0.351 | 0.94 [0.84, 1.04] | $2.29\times10^{-1}$ | 0.350 | 0.92 [0.83, 1.01] | $7.61\times10^{-2}$ |
| [9] | rs518425 | 15:78883813 | *CHRNA5* | G | 0.272 | 1.15 [0.92, 1.15] | $6.12\times10^{-1}$ | 0.271 | 1.07 [0.97, 1.19] | $1.71\times10^{-1}$ |
| [11] | rs8109525 | 19:41491918 | *CYP2B6* | G | 0.163 | 1.11 [0.90, 1.11] | $9.71\times10^{-1}$ | 0.162 | 1.01 [0.92, 1.12] | $7.76\times10^{-1}$ |
| [9] | rs2236196 | 20:61977556 | *CHRNA4* | G | 0.339 | 1.07 [0.85, 1.07] | $4.31\times10^{-1}$ | 0.340 | 0.86 [0.77, 0.96] | $5.29\times10^{-3}$ |
| [9] | rs3787138 | 20:61979224 | *CHRNA4* | G | 0.253 | 1.09 [0.80, 1.09] | $3.83\times10^{-1}$ | 0.252 | 0.83 [0.71, 0.95] | $9.18\times10^{-3}$ |
| [9] | rs6062899 | 20:61979793 | *CHRNA4* | G | 0.124 | 1.11 [0.86, 1.11] | $7.31\times10^{-1}$ | 0.121 | 0.85 [0.75, 0.96] | $1.09\times10^{-2}$ |
| [12] | rs1044396 | 20:61981134 | *CHRNA4* | T | 0.175 | 1.13 [0.92, 1.13] | $6.69\times10^{-1}$ | 0.173 | 0.96 [0.88, 1.06] | $4.16\times10^{-1}$ |
| **Nausea severity post-varenicline treatment** | | | | | | | | | | |
| [13] | rs2072660 | 1:154548721 | *CHRNB2* | T | 0.241 | 1.04 [0.92, 1.16] | $5.58\times10^{-1}$ | 0.238 | 1.04 [0.93, 1.15] | $5.15\times10^{-1}$ |
| [13] | rs2072661 | 1:154548880 | *CHRNB2* | G | 0.756 | 0.96 [0.85, 1.08] | $4.67\times10^{-1}$ | 0.758 | 0.96 [0.87, 1.07] | $4.84\times10^{-1}$ |
| [13] | rs4292956 | 1:154548946 | *CHRNB2* | C | 0.925 | 1.03 [0.84, 1.26] | $7.59\times10^{-1}$ | 0.927 | 0.97 [0.80, 1.16] | $7.19\times10^{-1}$ |
| [13] | rs2302764 | 17:7360110 | *CHRNB1* | C | 0.163 | 0.94 [0.82, 1.08] | $3.77\times10^{-1}$ | 0.162 | 1.02 [0.89, 1.15] | $8.05\times10^{-1}$ |
| **Smoking cessation** | | | | | | | | | | |
| [14] | rs7521775 | 1:22475649 | Intergenic | C | 0.590 | 1.12 [1.01, 1.24] | $2.69\times10^{-2}$ | 0.587 | 1.06 [0.97, 1.16] | $2.27\times10^{-1}$ |
| [14] | rs10794514 | 1:28659696 | *MED18* | T | 0.427 | 1.00 [0.91, 1.11] | $9.63\times10^{-1}$ | 0.427 | 1.00 [0.91, 1.10] | $9.85\times10^{-1}$ |
| [14] | rs3791151 | 1:44078602 | *PTPRF* | C | 0.662 | 0.99 [0.89, 1.10] | $8.34\times10^{-1}$ | 0.665 | 0.97 [0.88, 1.07] | $4.96\times10^{-1}$ |
| [14] | rs1171279 | 1:65988493 | *LEPR* | T | 0.271 | 1.03 [0.92, 1.15] | $6.32\times10^{-1}$ | 0.265 | 1.00 [0.90, 1.11] | $9.65\times10^{-1}$ |
| [14] | rs6677439 | 1:73837907 | Intergenic | C | 0.548 | 1.02 [0.91, 1.14] | $7.48\times10^{-1}$ | 0.549 | 1.02 [0.92, 1.14] | $6.80\times10^{-1}$ |
| [14] | rs699536 | 1:90934652 | Intergenic | G | 0.569 | 1.01 [0.92, 1.12] | $8.09\times10^{-1}$ | 0.575 | 0.99 [0.91, 1.09] | $9.09\times10^{-1}$ |
| [14] | rs17379561 | 1:98340139 | *DPYD* | T | 0.144 | 1.02 [0.88, 1.18] | $7.93\times10^{-1}$ | 0.148 | 1.10 [0.97, 1.25] | $1.53\times10^{-1}$ |

| | | | | | | | | | | |
|---|---|---|---|---|---|---|---|---|---|---|
| 14 | rs12727810 | 1:162068428 | *NOS1AP* | G | 0.371 | 1.01 [0.91, 1.13] | $7.87\times10^{-1}$ | 0.371 | 1.01 [0.92, 1.11] | $8.39\times10^{-1}$ |
| 14 | rs9724968 | 1:162069609 | *NOS1AP* | T | 0.374 | 1.00 [0.90, 1.12] | $9.88\times10^{-1}$ | 0.376 | 1.01 [0.91, 1.12] | $8.70\times10^{-1}$ |
| 14 | rs6673326 | 1:169420973 | *CCDC181* | G | 0.375 | 1.09 [0.98, 1.22] | $1.20\times10^{-1}$ | 0.373 | 1.00 [0.90, 1.11] | $9.99\times10^{-1}$ |
| 14 | rs6425284 | 1:174499977 | *RABGAP1L* | G | 0.707 | 1.11 [0.98, 1.27] | $1.12\times10^{-1}$ | 0.706 | 1.03 [0.91, 1.17] | $6.09\times10^{-1}$ |
| 14 | rs12046747 | 1:204593696 | *LRRN2* | G | 0.792 | 0.93 [0.82, 1.05] | $2.19\times10^{-1}$ | 0.792 | 1.05 [0.93, 1.17] | $4.47\times10^{-1}$ |
| 14 | rs72781639 | 2:24204148 | *UBXN2A* | G | 0.156 | 1.01 [0.87, 1.16] | $9.30\times10^{-1}$ | 0.154 | 1.04 [0.91, 1.18] | $5.94\times10^{-1}$ |
| 14 | rs12614418 | 2:35730212 | Intergenic | G | 0.442 | 1.02 [0.91, 1.14] | $7.34\times10^{-1}$ | 0.439 | 1.03 [0.93, 1.14] | $5.57\times10^{-1}$ |
| 14 | rs7569203 | 2:45154418 | *LINC01833* | C | 0.318 | 0.89 [0.80, 0.99] | $3.26\times10^{-2}$ | 0.325 | 0.94 [0.85, 1.04] | $2.34\times10^{-1}$ |
| 14 | rs1369294 | 2:50232133 | *NRXN1* | T | 0.472 | 0.97 [0.88, 1.07] | $5.98\times10^{-1}$ | 0.470 | 0.98 [0.90, 1.08] | $7.25\times10^{-1}$ |
| 14 | rs10191428 | 2:62725407 | Intergenic | C | 0.903 | 1.05 [0.88, 1.24] | $6.12\times10^{-1}$ | 0.903 | 0.95 [0.81, 1.11] | $5.28\times10^{-1}$ |
| 14 | rs7598901 | 2:73675844 | *ALMS1* | C | 0.384 | 1.04 [0.94, 1.16] | $4.36\times10^{-1}$ | 0.380 | 1.05 [0.95, 1.15] | $3.35\times10^{-1}$ |
| 14 | rs2916251 | 2:104447052 | Intergenic | T | 0.510 | 1.03 [0.92, 1.16] | $5.90\times10^{-1}$ | 0.506 | 1.04 [0.93, 1.15] | $5.04\times10^{-1}$ |
| 14 | rs1374328 | 2:166182354 | *SCN2A* | T | 0.756 | 0.97 [0.86, 1.09] | $5.90\times10^{-1}$ | 0.754 | 1.04 [0.94, 1.16] | $4.39\times10^{-1}$ |
| 14 | rs78590213 | 2:181920762 | *UBE2E3* | T | 0.707 | 1.05 [0.94, 1.17] | $3.96\times10^{-1}$ | 0.711 | 1.00 [0.90, 1.11] | $9.77\times10^{-1}$ |
| 14 | rs56222728 | 2:186135147 | Intergenic | T | 0.610 | 1.06 [0.95, 1.18] | $3.19\times10^{-1}$ | 0.607 | 1.12 [1.01, 1.24] | $3.49\times10^{-2}$ |
| 14 | rs12693975 | 2:203720745 | *ICA1L* | A | 0.822 | 1.02 [0.90, 1.17] | $7.16\times10^{-1}$ | 0.826 | 1.02 [0.90, 1.15] | $7.78\times10^{-1}$ |
| 14 | rs34762726 | 3:49689210 | *BSN* | G | 0.720 | 0.91 [0.81, 1.02] | $9.32\times10^{-2}$ | 0.723 | 0.94 [0.85, 1.04] | $2.16\times10^{-1}$ |
| 14 | rs7613360 | 3:49916710 | Intergenic | T | 0.411 | 0.95 [0.86, 1.05] | $3.13\times10^{-1}$ | 0.414 | 0.95 [0.87, 1.05] | $3.25\times10^{-1}$ |
| 14 | rs6803651 | 3:64431730 | Intergenic | G | 0.565 | 0.99 [0.90, 1.10] | $8.69\times10^{-1}$ | 0.566 | 0.93 [0.85, 1.02] | $1.31\times10^{-1}$ |
| 14 | rs9833290 | 3:77312267 | *ROBO2* | C | 0.402 | 0.98 [0.88, 1.08] | $6.61\times10^{-1}$ | 0.408 | 0.98 [0.89, 1.08] | $6.79\times10^{-1}$ |
| 14 | rs2171141 | 3:85177049 | *CADM2* | C | 0.437 | 1.07 [0.97, 1.18] | $1.86\times10^{-1}$ | 0.439 | 1.06 [0.97, 1.16] | $2.10\times10^{-1}$ |
| 14 | rs1870008 | 3:107954891 | Intergenic | T | 0.605 | 0.94 [0.85, 1.04] | $2.36\times10^{-1}$ | 0.601 | 0.97 [0.89, 1.07] | $5.95\times10^{-1}$ |
| 14 | rs1711161 | 3:136080537 | *STAG1* | T | 0.749 | 1.06 [0.95, 1.19] | $3.12\times10^{-1}$ | 0.749 | 1.08 [0.97, 1.20] | $1.80\times10^{-1}$ |
| 14 | rs36035502 | 4:35580129 | Intergenic | C | 0.194 | 0.96 [0.84, 1.10] | $5.42\times10^{-1}$ | 0.196 | 0.84 [0.74, 0.96] | $8.65\times10^{-3}$ |
| 14 | rs75096215 | 4:35582194 | Intergenic | G | 0.195 | 0.93 [0.82, 1.05] | $2.41\times10^{-1}$ | 0.198 | 0.86 [0.76, 0.97] | $1.24\times10^{-2}$ |
| 14 | rs76757555 | 4:35582979 | Intergenic | A | 0.195 | 0.93 [0.82, 1.05] | $2.38\times10^{-1}$ | 0.198 | 0.86 [0.76, 0.97] | $1.26\times10^{-2}$ |
| 14 | rs77698860 | 4:35582980 | Intergenic | T | 0.195 | 0.93 [0.82, 1.05] | $2.40\times10^{-1}$ | 0.198 | 0.86 [0.76, 0.97] | $1.26\times10^{-2}$ |
| 14 | rs11937919 | 4:35583243 | Intergenic | G | 0.196 | 0.93 [0.82, 1.05] | $2.43\times10^{-1}$ | 0.199 | 0.86 [0.76, 0.97] | $1.20\times10^{-2}$ |
| 14 | rs115737563 | 4:42140389 | *BEND4* | A | 0.197 | 1.12 [0.98, 1.28] | $8.33\times10^{-2}$ | 0.194 | 1.06 [0.93, 1.19] | $3.80\times10^{-1}$ |
| 14 | rs62310097 | 4:93499316 | *GRID2* | T | 0.362 | 0.99 [0.88, 1.11] | $8.35\times10^{-1}$ | 0.362 | 0.95 [0.86, 1.06] | $3.80\times10^{-1}$ |
| 14 | rs11721364 | 4:105427978 | *CXXC4-AS1* | C | 0.679 | 1.04 [0.93, 1.15] | $5.20\times10^{-1}$ | 0.675 | 1.00 [0.91, 1.10] | $9.90\times10^{-1}$ |

| | | | | | | | | | | |
|---|---|---|---|---|---|---|---|---|---|---|
| 14 | rs1000350 | 4:113044466 | Intergenic | T | 0.375 | 0.99 [0.89, 1.10] | $8.69 \times 10^{-1}$ | 0.378 | 1.01 [0.91, 1.12] | $8.61 \times 10^{-1}$ |
| 14 | rs55669488 | 4:140943275 | *MAML3* | T | 0.642 | 1.03 [0.93, 1.14] | $6.09 \times 10^{-1}$ | 0.638 | 1.03 [0.93, 1.13] | $6.05 \times 10^{-1}$ |
| 14 | rs7668995 | 4:147919278 | Intergenic | T | 0.720 | 1.09 [0.97, 1.21] | $1.43 \times 10^{-1}$ | 0.722 | 0.97 [0.87, 1.07] | $5.36 \times 10^{-1}$ |
| 14 | rs4691521 | 4:159802373 | *FNIP2* | G | 0.431 | 1.00 [0.89, 1.11] | $9.45 \times 10^{-1}$ | 0.431 | 1.02 [0.92, 1.13] | $6.65 \times 10^{-1}$ |
| 14 | rs7705852 | 5:59855859 | Intergenic | T | 0.165 | 1.02 [0.89, 1.17] | $7.35 \times 10^{-1}$ | 0.168 | 1.12 [0.99, 1.26] | $8.49 \times 10^{-2}$ |
| 14 | rs329120 | 5:133861756 | *JADE2* | C | 0.581 | 1.05 [0.95, 1.16] | $3.24 \times 10^{-1}$ | 0.581 | 1.05 [0.96, 1.15] | $2.92 \times 10^{-1}$ |
| 14 | rs4705014 | 5:155852315 | *SGCD* | G | 0.377 | 1.03 [0.93, 1.14] | $6.02 \times 10^{-1}$ | 0.374 | 1.08 [0.98, 1.19] | $1.02 \times 10^{-1}$ |
| 14 | rs263886 | 5:165170651 | Intergenic | A | 0.509 | 0.98 [0.88, 1.08] | $6.34 \times 10^{-1}$ | 0.506 | 0.96 [0.88, 1.06] | $4.49 \times 10^{-1}$ |
| 14 | rs11738110 | 5:166992760 | *TENM2* | T | 0.378 | 0.97 [0.87, 1.07] | $5.18 \times 10^{-1}$ | 0.373 | 1.04 [0.95, 1.15] | $3.86 \times 10^{-1}$ |
| 14 | rs2073526 | 6:26374658 | *BTN3A2* | G | 0.454 | 0.93 [0.84, 1.02] | $1.32 \times 10^{-1}$ | 0.457 | 0.92 [0.84, 1.01] | $8.35 \times 10^{-2}$ |
| 14 | rs13203795 | 6:33583732 | Intergenic | A | 0.100 | 1.07 [0.89, 1.28] | $4.81 \times 10^{-1}$ | 0.102 | 1.05 [0.88, 1.24] | $5.88 \times 10^{-1}$ |
| 14 | rs7778443 | 7:32314690 | *PDE1C* | T | 0.403 | 1.11 [1.01, 1.23] | $3.93 \times 10^{-2}$ | 0.397 | 1.07 [0.98, 1.18] | $1.39 \times 10^{-1}$ |
| 14 | rs62459050 | 7:71319137 | *CALN1* | G | 0.360 | 0.89 [0.80, 0.99] | $3.52 \times 10^{-2}$ | 0.363 | 0.95 [0.86, 1.05] | $2.98 \times 10^{-1}$ |
| 14 | rs113067414 | 7:97391962 | Intergenic | G | 0.758 | 0.98 [0.87, 1.10] | $7.18 \times 10^{-1}$ | 0.757 | 0.97 [0.87, 1.09] | $6.41 \times 10^{-1}$ |
| 14 | rs13245429 | 7:97394886 | Intergenic | G | 0.795 | 0.96 [0.84, 1.10] | $5.30 \times 10^{-1}$ | 0.796 | 0.94 [0.83, 1.07] | $3.60 \times 10^{-1}$ |
| 14 | rs7794285 | 7:104849737 | *SRPK2* | A | 0.451 | 0.95 [0.86, 1.05] | $2.78 \times 10^{-1}$ | 0.456 | 1.00 [0.91, 1.09] | $9.47 \times 10^{-1}$ |
| 14 | rs4265129 | 7:113591217 | Intergenic | A | 0.373 | 1.16 [1.04, 1.28] | $6.15 \times 10^{-3}$ | 0.372 | 1.02 [0.93, 1.13] | $6.45 \times 10^{-1}$ |
| 14 | rs62474673 | 7:114973194 | Intergenic | T | 0.499 | 0.98 [0.89, 1.08] | $7.21 \times 10^{-1}$ | 0.499 | 1.01 [0.92, 1.10] | $9.15 \times 10^{-1}$ |
| 14 | rs12154872 | 7:117512932 | *CTTNBP2* | G | 0.555 | 0.96 [0.86, 1.06] | $3.89 \times 10^{-1}$ | 0.560 | 0.97 [0.88, 1.06] | $4.58 \times 10^{-1}$ |
| 14 | rs2098112 | 7:153487944 | *DPP6* | A | 0.482 | 0.91 [0.83, 1.01] | $7.22 \times 10^{-2}$ | 0.482 | 0.99 [0.91, 1.09] | $8.73 \times 10^{-1}$ |
| 14 | rs3807890 | 7:155571603 | *RBM33* | G | 0.198 | 1.01 [0.89, 1.15] | $8.58 \times 10^{-1}$ | 0.199 | 0.97 [0.86, 1.10] | $6.35 \times 10^{-1}$ |
| 14 | rs11991338 | 8:9288464 | Intergenic | A | 0.178 | 0.91 [0.79, 1.05] | $1.82 \times 10^{-1}$ | 0.177 | 0.99 [0.87, 1.13] | $8.74 \times 10^{-1}$ |
| 14 | rs2736268 | 8:11188532 | *SLC35G5* | C | 0.534 | 0.98 [0.88, 1.09] | $6.91 \times 10^{-1}$ | 0.530 | 0.98 [0.89, 1.09] | $7.65 \times 10^{-1}$ |
| 14 | rs11773992 | 8:12667804 | *LINC00681\|LOC340357* | C | 0.188 | 0.92 [0.81, 1.05] | $2.18 \times 10^{-1}$ | 0.188 | 0.95 [0.84, 1.07] | $3.82 \times 10^{-1}$ |
| 14 | rs11776293 | 8:27418429 | Intergenic | C | 0.834 | 0.98 [0.85, 1.12] | $7.40 \times 10^{-1}$ | 0.833 | 0.91 [0.80, 1.03] | $1.30 \times 10^{-1}$ |
| 14 | rs1565735 | 8:27426077 | Intergenic | T | 0.812 | 0.92 [0.81, 1.04] | $1.91 \times 10^{-1}$ | 0.810 | 0.94 [0.83, 1.06] | $2.91 \times 10^{-1}$ |
| 14 | rs11990422 | 8:42582955 | *CHRNB3* | G | 0.897 | 1.00 [0.84, 1.17] | $9.56 \times 10^{-1}$ | 0.898 | 1.06 [0.91, 1.24] | $4.69 \times 10^{-1}$ |
| 14 | rs10738785 | 9:27919960 | Intergenic | A | 0.350 | 0.96 [0.86, 1.06] | $3.99 \times 10^{-1}$ | 0.355 | 0.93 [0.85, 1.03] | $1.71 \times 10^{-1}$ |
| 14 | rs10819036 | 9:127890913 | *SCAI* | T | 0.700 | 0.98 [0.88, 1.10] | $7.52 \times 10^{-1}$ | 0.700 | 1.01 [0.91, 1.11] | $8.87 \times 10^{-1}$ |
| 14 | rs4837011 | 9:127923014 | *PPP6C* | G | 0.699 | 0.93 [0.82, 1.04] | $1.96 \times 10^{-1}$ | 0.699 | 0.96 [0.86, 1.07] | $4.48 \times 10^{-1}$ |
| 14 | rs4838291 | 9:128489937 | Intergenic | G | 0.392 | 1.08 [0.97, 1.19] | $1.65 \times 10^{-1}$ | 0.391 | 1.01 [0.92, 1.11] | $8.66 \times 10^{-1}$ |

| | | | | | | | | | | |
|---|---|---|---|---|---|---|---|---|---|---|
| [14] | rs112883710 | 9:136430088 | *ADAMTSL2* | A | 0.110 | 0.99 [0.83, 1.17] | $8.85\times10^{-1}$ | 0.110 | 0.95 [0.80, 1.11] | $5.02\times10^{-1}$ |
| [14] | rs112395992 | 9:136449214 | Intergenic | T | 0.042 | 1.01 [0.66, 1.54] | $9.75\times10^{-1}$ | 0.056 | 0.95 [0.65, 1.40] | $8.09\times10^{-1}$ |
| [14] | rs113067637 | 9:136452411 | Intergenic | A | 0.121 | 1.02 [0.87, 1.19] | $8.33\times10^{-1}$ | 0.120 | 1.02 [0.88, 1.18] | $7.76\times10^{-1}$ |
| [14] | rs3888560 | 9:136465342 | Intergenic | T | 0.301 | 1.00 [0.89, 1.13] | $9.55\times10^{-1}$ | 0.302 | 1.03 [0.92, 1.15] | $6.13\times10^{-1}$ |
| [14] | rs78094364 | 9:136466679 | Intergenic | T | 0.963 | 0.76 [0.58, 1.00] | $5.19\times10^{-2}$ | 0.957 | 0.84 [0.66, 1.07] | $1.61\times10^{-1}$ |
| [14] | rs739447 | 9:136477511 | Intergenic | T | 0.135 | 1.02 [0.88, 1.18] | $8.01\times10^{-1}$ | 0.132 | 0.96 [0.84, 1.11] | $6.06\times10^{-1}$ |
| [14] | rs7872903 | 9:136484292 | Intergenic | C | 0.251 | 1.04 [0.93, 1.17] | $4.87\times10^{-1}$ | 0.251 | 0.94 [0.84, 1.04] | $2.17\times10^{-1}$ |
| [14] | rs1108581 | 9:136505241 | *DBH* | A | 0.809 | 1.08 [0.95, 1.23] | $2.28\times10^{-1}$ | 0.808 | 0.95 [0.85, 1.07] | $4.02\times10^{-1}$ |
| [14] | rs10509123 | 10:61893145 | *ANK3* | T | 0.635 | 1.00 [0.90, 1.11] | $1.00\times10^{0}$ | 0.636 | 0.91 [0.83, 1.00] | $6.05\times10^{-2}$ |
| [14] | rs7896518 | 10:65104500 | *JMJD1C* | A | 0.581 | 1.00 [0.91, 1.11] | $9.46\times10^{-1}$ | 0.582 | 1.01 [0.92, 1.11] | $8.66\times10^{-1}$ |
| [14] | rs35649565 | 10:87074564 | Intergenic | G | 0.759 | 0.95 [0.85, 1.07] | $3.93\times10^{-1}$ | 0.761 | 1.06 [0.96, 1.19] | $2.58\times10^{-1}$ |
| [14] | rs61861137 | 10:105613383 | *SH3PXD2A* | A | 0.759 | 1.01 [0.90, 1.13] | $8.81\times10^{-1}$ | 0.753 | 0.94 [0.84, 1.05] | $2.50\times10^{-1}$ |
| [14] | rs1827535 | 11:16380409 | *SOX6* | G | 0.329 | 1.03 [0.92, 1.15] | $5.93\times10^{-1}$ | 0.328 | 1.00 [0.90, 1.10] | $9.53\times10^{-1}$ |
| [14] | rs2585817 | 11:28602221 | Intergenic | A | 0.633 | 0.99 [0.90, 1.10] | $8.82\times10^{-1}$ | 0.631 | 1.07 [0.97, 1.18] | $1.61\times10^{-1}$ |
| [14] | rs1783818 | 11:57437551 | ZDHHC5 | T | 0.562 | 1.04 [0.94, 1.15] | $4.57\times10^{-1}$ | 0.563 | 1.04 [0.94, 1.14] | $4.64\times10^{-1}$ |
| [14] | rs10792430 | 11:64147114 | Intergenic | T | 0.816 | 0.97 [0.86, 1.11] | $6.78\times10^{-1}$ | 0.815 | 1.04 [0.92, 1.17] | $5.48\times10^{-1}$ |
| [14] | rs2734838 | 11:113286501 | *DRD2* | A | 0.407 | 1.05 [0.95, 1.17] | $3.09\times10^{-1}$ | 0.403 | 1.00 [0.91, 1.10] | $9.84\times10^{-1}$ |
| [14] | rs11223560 | 11:133561399 | Intergenic | G | 0.418 | 0.98 [0.88, 1.08] | $6.68\times10^{-1}$ | 0.418 | 0.97 [0.88, 1.06] | $4.86\times10^{-1}$ |
| [14] | rs11171710 | 12:56368078 | *RAB5B* | A | 0.459 | 1.08 [0.98, 1.19] | $1.23\times10^{-1}$ | 0.459 | 0.97 [0.88, 1.06] | $4.75\times10^{-1}$ |
| [14] | rs4417327 | 12:58285460 | *LOC283387* | C | 0.563 | 0.96 [0.87, 1.06] | $4.15\times10^{-1}$ | 0.563 | 0.97 [0.89, 1.07] | $5.71\times10^{-1}$ |
| [14] | rs8756 | 12:66359752 | *HMGA2* | A | 0.512 | 1.14 [1.03, 1.26] | $1.12\times10^{-2}$ | 0.505 | 1.14 [1.04, 1.25] | $5.61\times10^{-3}$ |
| [14] | rs17113564 | 12:74904625 | Intergenic | C | 0.867 | 1.04 [0.90, 1.21] | $5.68\times10^{-1}$ | 0.868 | 1.04 [0.91, 1.19] | $5.92\times10^{-1}$ |
| [14] | rs7960340 | 12:99737259 | *ANKS1B* | A | 0.142 | 0.92 [0.80, 1.07] | $2.91\times10^{-1}$ | 0.142 | 0.94 [0.82, 1.07] | $3.48\times10^{-1}$ |
| [14] | rs678436 | 12:111949394 | *ATXN2* | C | 0.256 | 0.96 [0.86, 1.08] | $5.42\times10^{-1}$ | 0.259 | 1.00 [0.89, 1.11] | $9.32\times10^{-1}$ |
| [14] | rs4902704 | 14:69703588 | *EXD2* | C | 0.624 | 0.91 [0.82, 1.00] | $6.18\times10^{-2}$ | 0.625 | 0.94 [0.85, 1.03] | $1.80\times10^{-1}$ |
| [14] | rs12884296 | 14:79534362 | *NRXN3* | A | 0.386 | 1.01 [0.91, 1.12] | $8.50\times10^{-1}$ | 0.383 | 1.02 [0.92, 1.12] | $7.28\times10^{-1}$ |
| [14] | rs7152530 | 14:98641215 | Intergenic | A | 0.363 | 0.96 [0.86, 1.06] | $4.00\times10^{-1}$ | 0.362 | 0.96 [0.88, 1.06] | $4.43\times10^{-1}$ |
| [14] | rs34161718 | 14:104620193 | *KIF26A* | C | 0.772 | 0.98 [0.87, 1.10] | $7.22\times10^{-1}$ | 0.768 | 1.02 [0.92, 1.15] | $6.82\times10^{-1}$ |
| [14] | rs35534970 | 15:47675585 | *SEMA6D* | A | 0.225 | 1.02 [0.91, 1.15] | $7.43\times10^{-1}$ | 0.228 | 1.09 [0.98, 1.22] | $1.03\times10^{-1}$ |
| [14] | rs58365910 | 15:78849034 | Intergenic | C | 0.354 | 0.92 [0.83, 1.02] | $1.17\times10^{-1}$ | 0.352 | 0.91 [0.82, 1.00] | $5.38\times10^{-2}$ |
| [14] | NA | 15:78850501 | Intergenic | A | 0.353 | 0.94 [0.84, 1.05] | $2.67\times10^{-1}$ | 0.349 | 0.90 [0.81, 1.00] | $4.33\times10^{-2}$ |

| | | | | | | | | | | |
|---|---|---|---|---|---|---|---|---|---|---|
| [14] | rs667282 | 15:78863472 | *CHRNA5* | T | 0.790 | 0.98 [0.87, 1.11] | $7.57\times10^{-1}$ | 0.790 | 0.91 [0.81, 1.02] | $9.24\times10^{-2}$ |
| [14] | rs150300 | 15:89944190 | Intergenic | T | 0.606 | 0.94 [0.84, 1.05] | $2.97\times10^{-1}$ | 0.608 | 0.94 [0.84, 1.04] | $2.48\times10^{-1}$ |
| [14] | rs35198836 | 16:1244631 | *CACNA1H* | T | 0.304 | 0.97 [0.87, 1.08] | $5.55\times10^{-1}$ | 0.303 | 0.94 [0.85, 1.04] | $2.50\times10^{-1}$ |
| [14] | rs75744751 | 16:53564401 | Intergenic | C | 0.061 | 1.06 [0.86, 1.31] | $5.92\times10^{-1}$ | 0.061 | 1.11 [0.91, 1.35] | $2.96\times10^{-1}$ |
| [14] | rs1364349 | 16:61029491 | Intergenic | C | 0.907 | 0.97 [0.81, 1.15] | $7.13\times10^{-1}$ | 0.905 | 0.88 [0.75, 1.02] | $9.70\times10^{-2}$ |
| [14] | rs3759982 | 16:69140747 | *HAS3* | C | 0.126 | 1.05 [0.90, 1.21] | $5.60\times10^{-1}$ | 0.127 | 0.93 [0.81, 1.07] | $3.34\times10^{-1}$ |
| [14] | rs727428 | 17:7537792 | Intergenic | C | 0.556 | 0.92 [0.83, 1.02] | $1.28\times10^{-1}$ | 0.559 | 0.97 [0.88, 1.06] | $4.82\times10^{-1}$ |
| [14] | rs4925130 | 17:17807858 | *TOM1L2* | A | 0.372 | 1.01 [0.91, 1.11] | $9.14\times10^{-1}$ | 0.372 | 0.96 [0.87, 1.05] | $3.81\times10^{-1}$ |
| [14] | rs66481362 | 17:37758172 | Intergenic | A | 0.085 | 1.03 [0.86, 1.24] | $7.54\times10^{-1}$ | 0.083 | 0.95 [0.80, 1.13] | $5.74\times10^{-1}$ |
| [14] | rs1941472 | 18:37702403 | Intergenic | G | 0.438 | 1.00 [0.90, 1.11] | $9.87\times10^{-1}$ | 0.439 | 0.95 [0.86, 1.05] | $3.15\times10^{-1}$ |
| [14] | rs61537869 | 18:50542722 | *DCC* | G | 0.589 | 0.96 [0.87, 1.07] | $4.56\times10^{-1}$ | 0.588 | 0.99 [0.90, 1.09] | $8.77\times10^{-1}$ |
| [14] | rs11881918 | 19:41334199 | Intergenic | A | 0.072 | 1.1 [0.90, 1.35] | $3.34\times10^{-1}$ | 0.074 | 1.00 [0.83, 1.20] | $9.64\times10^{-1}$ |
| [14] | rs10853742 | 19:41340573 | Intergenic | G | 0.345 | 0.95 [0.85, 1.05] | $3.23\times10^{-1}$ | 0.347 | 0.97 [0.87, 1.07] | $4.88\times10^{-1}$ |
| [14] | rs56113850 | 19:41353107 | *CYP2A6* | T | 0.432 | 0.91 [0.82, 1.01] | $6.53\times10^{-2}$ | 0.432 | 0.93 [0.85, 1.02] | $1.41\times10^{-1}$ |
| [14] | rs28399443 | 19:41354306 | *CYP2A6* | A | 0.022 | 0.77 [0.53, 1.12] | $1.73\times10^{-1}$ | 0.023 | 1.11 [0.80, 1.54] | $5.35\times10^{-1}$ |
| [14] | rs57837628 | 19:41357910 | Intergenic | A | 0.482 | 0.92 [0.83, 1.01] | $9.05\times10^{-2}$ | 0.481 | 0.91 [0.83, 1.00] | $4.18\times10^{-2}$ |
| [14] | rs117824460 | 19:41371480 | Intergenic | G | 0.031 | 1.12 [0.81, 1.54] | $4.82\times10^{-1}$ | 0.031 | 1.21 [0.89, 1.63] | $2.19\times10^{-1}$ |
| [14] | rs6011779 | 20:61984317 | *CHRNA4* | C | 0.208 | 1.02 [0.91, 1.16] | $7.19\times10^{-1}$ | 0.206 | 0.90 [0.80, 1.01] | $7.23\times10^{-2}$ |
| [14] | rs12479680 | 20:61988413 | *CHRNA4* | A | 0.215 | 1.00 [0.88, 1.14] | $9.86\times10^{-1}$ | 0.213 | 0.91 [0.80, 1.03] | $1.29\times10^{-1}$ |
| [14] | rs45497800 | 20:61991833 | *CHRNA4*\|*LOC100130587* | T | 0.093 | 1.15 [0.97, 1.37] | $1.13\times10^{-1}$ | 0.095 | 1.09 [0.93, 1.27] | $3.12\times10^{-1}$ |
| [14] | rs79739740 | 20:61992467 | *CHRNA4* | T | 0.089 | 1.15 [0.94, 1.41] | $1.84\times10^{-1}$ | 0.089 | 1.09 [0.91, 1.32] | $3.48\times10^{-1}$ |
| [14] | rs11699828 | 20:62157198 | Intergenic | G | 0.970 | 1.09 [0.77, 1.55] | $6.31\times10^{-1}$ | 0.970 | 1.32 [0.96, 1.83] | $9.23\times10^{-2}$ |
| [14] | rs9611540 | 22:41674203 | *RANGAP1* | C | 0.673 | 1.01 [0.91, 1.12] | $8.83\times10^{-1}$ | 0.675 | 0.94 [0.85, 1.04] | $2.50\times10^{-1}$ |
| [14] | rs2542028 | 22:47196524 | *TBC1D22A* | A | 0.277 | 1.08 [0.96, 1.20] | $1.93\times10^{-1}$ | 0.282 | 1.06 [0.96, 1.18] | $2.43\times10^{-1}$ |

Odds ratios (ORs) and 95% confidence intervals (CIs) are aligned to the allele (EA) which increases risk of varenicline efficacy, nausea and smoking cessation.
*Abbreviations:* Chr, chromosome; EAF, effect allele frequency.

**Supplementary Table 6** Look-up of sentinel variants associated with **(A)** SVE and **(B)** LVE in genome-wide association studies of age at smoking initiation, smoking initiation, cigarettes per day and smoking cessation performed by GSCAN[14].

| Sentinel variant | Chr:Position (hg19) | EA | Age at initiation | | Smoking initiation | | Cigarettes per day | | Smoking cessation | |
|---|---|---|---|---|---|---|---|---|---|---|
| | | | **OR [95% CI]** | ***p*-value** | **OR [95% CI]** | ***p*-value** | **OR [95% CI]** | ***p*-value** | **OR [95% CI]** | ***p*-value** |
| **(A) Short-term varenicline effectiveness (SVE)** | | | | | | | | | | |
| rs186444103 | 2:40054022 | T | 0.98 [0.96, 1.00] | 2.61E-02 | 0.99 [0.98, 1.00] | 9.33E-02 | 0.99 [0.97, 1.01] | 3.47E-01 | 1.00 [0.98, 1.01] | 8.41E-01 |
| rs4364036 | 2:162904624 | T | 1.00 [0.98, 1.01] | 8.56E-01 | 1.00 [0.99, 1.01] | 9.48E-01 | 1.01 [0.99, 1.03] | 2.12E-01 | 1.00 [0.99, 1.02] | 7.38E-01 |
| rs4686373 | 3:9853437 | A | 1.00 [0.99, 1.01] | 7.97E-01 | 1.00 [0.99, 1.01] | 9.41E-01 | 1.00 [0.99, 1.01] | 3.58E-01 | 1.00 [0.99, 1.01] | 6.68E-01 |
| rs78597169 | 8:117830288 | A | 0.99 [0.98, 1.01] | 4.75E-01 | 1.01 [1.00, 1.03] | 1.67E-02 | 1.00 [0.98, 1.02] | 8.68E-01 | 1.00 [0.98, 1.01] | 8.01E-01 |
| rs2297038 | 10:30663464 | T | 1.00 [0.99, 1.01] | 8.49E-01 | 1.00 [1.00, 1.01] | 4.52E-01 | 1.00 [0.99, 1.01] | 8.85E-01 | 1.01 [1.00, 1.01] | 4.72E-02 |
| rs9600669 | 13:76776621 | A | 1.00 [0.99, 1.01] | 6.45E-01 | 1.00 [0.99, 1.00] | 3.68E-01 | 0.99 [0.98, 1.00] | 5.18E-02 | 1.01 [1.00, 1.02] | 1.36E-02 |
| rs8003262 | 14:102050501 | A | 1.01 [1.00, 1.02] | 1.67E-02 | 1.00 [0.99, 1.01] | 8.88E-01 | 1.00 [0.99, 1.01] | 4.51E-01 | 1.00 [0.99, 1.01] | 8.92E-01 |
| rs827937 | 20:46793101 | A | 1.00 [0.99, 1.00] | 2.81E-01 | 1.00 [0.99, 1.01] | 7.90E-01 | 1.00 [0.99, 1.00] | 1.59E-01 | 1.00 [0.99, 1.02] | 6.36E-01 |
| **(B) Long-term varenicline effectiveness (LVE)** | | | | | | | | | | |
| rs895545 | 2:121721467 | T | 1.00 [0.99, 1.01] | 7.16E-01 | 1.00 [0.99, 1.01] | 8.61E-01 | 0.99 [0.98, 1.00] | 2.36E-01 | 1.01 [1.00, 1.02] | 1.66E-01 |
| rs7712227 | 5:118656769 | A | 0.99 [0.98, 1.00] | 1.11E-01 | 1.00 [0.99, 1.00] | 2.14E-01 | 1.01 [1.00, 1.02] | 1.36E-02 | 1.00 [0.99, 1.01] | 4.10E-01 |
| rs78695099 | 6:100195347 | A | 1.02 [1.00, 1.03] | 5.87E-02 | 0.98 [0.97, 0.99] | 1.68E-03 | 1.01 [0.99, 1.02] | 4.71E-01 | 1.00 [0.98, 1.02] | 9.77E-01 |
| rs35443123 | 7:85173800 | A | 1.00 [0.99, 1.00] | 3.44E-01 | 1.00 [1.00, 1.01] | 2.36E-02 | 1.00 [0.99, 1.00] | 1.72E-01 | 1.00 [0.99, 1.00] | 1.15E-01 |
| rs766169 | 22:47680100 | T | 1.00 [0.99, 1.01] | 5.47E-01 | 1.01 [1.00, 1.02] | 1.67E-02 | 1.00 [0.99, 1.01] | 6.84E-01 | 1.00 [0.99, 1.01] | 6.43E-01 |

SVE sentinels, rs149318872 and rs10421326, were not available in the GSCAN datasets and an appropriate proxy (within ±100kb with $r^2$ ≥0.9) could not be found. *Abbreviations:* Chr, chromosome; CI, confidence interval; EA, effect allele; OR, odds ratio.
